# Impacts of K-12 school reopening on the COVID-19 epidemic in Indiana, USA

**DOI:** 10.1101/2020.08.22.20179960

**Authors:** Guido España, Sean Cavany, Rachel Oidtman, Carly Barbera, Alan Costello, Anita Lerch, Marya Poterek, Quan Tran, Annaliese Wieler Sean Moore, T. Alex Perkins

## Abstract

In the United States, schools closed in March 2020 due to COVID-19 and began reopening in August 2020, despite continuing transmission of SARS-CoV-2. In states where in-person instruction resumed at that time, two major unknowns were the capacity at which schools would operate, which depended on the proportion of families opting for remote instruction, and adherence to face-mask requirements in schools, which depended on cooperation from students and enforcement by schools. To determine the impact of these conditions on the statewide burden of COVID-19 in Indiana, we used an agent-based model calibrated to and validated against multiple data types. Using this model, we quantified the burden of COVID-19 on K-12 students, teachers, their families, and the general population under alternative scenarios spanning three levels of school operating capacity (50%, 75%, and 100%) and three levels of face-mask adherence in schools (50%, 75%, and 100%). Under a scenario in which schools operated remotely, we projected 45,579 (95% CrI: 14,109-132,546) infections and 790 (95% CrI: 176-1680) deaths statewide between August 24 and December 31. Reopening at 100% capacity with 50% face-mask adherence in schools resulted in a proportional increase of 42.9 (95% CrI: 41.3-44.3) and 9.2 (95% CrI: 8.9-9.5) times that number of infections and deaths, respectively. In contrast, our results showed that at 50% capacity with 100% face-mask adherence, the number of infections and deaths were 22% (95% CrI: 16%-28%) and 11% (95% CrI: 5%-18%) higher than the scenario in which schools operated remotely. Within this range of possibilities, we found that high levels of school operating capacity (80-95%) and intermediate levels of face-mask adherence (40-70%) resulted in model behavior most consistent with observed data. Together, these results underscore the importance of precautions taken in schools for the benefit of their communities.

## 1 Introduction

The United States has been the country most severely impacted by the COVID-19 pandemic in terms of total reported cases and deaths, with over 28 million reported cases and more than 500 thousand deaths by March, 2021 [1]. This severity led to social interventions on an unprecedented scale, including restrictions on mass gatherings, bans on non-essential travel, and school closures [2, 3, 4, 5]. While such restrictions were initially successful in reducing transmission, the subsequent relaxation of restrictions on mass gatherings and movement were followed by large increases in notified cases and deaths [1, 3, 6, 7]. By the time the 2020-2021 school year began in August, transmission was at its highest point in the epidemic yet in some parts of the US. In Indiana, for example, the maximum number of daily cases was around 1,200 by then, which was higher than the previous maximum of fewer than 800 in late April [8].

This context of intense community transmission raised numerous questions about how schools should approach reopening for the start of the school year in August [9, 10, 11]. During influenza epidemics, school closures have been estimated to reduce transmission community-wide [12, 13, 14]. In general, schools are seen as key drivers of the transmission of respiratory pathogens due to close contact among children at school [15, 16, 17]. However, several factors complicated the effect of school reopenings on SARS-CoV-2 transmission. In particular, children and adolescents appear less susceptible to infection and are much less likely to experience severe outcomes following infection [18, 19, 20, 21, 22, 23]. It is also still unclear what their contribution to transmission is, but several studies suggest they can play an important role [18, 24, 25, 26]. There are additional economic and social factors to consider, too, such as the economic costs of school closures for families that must then stay home from work, and the nutritional benefits of school reopening for children who rely on free and subsidized school meals [27, 28, 29].

Our objective in this study was to explore how different conditions for school reopening during the fall semester of 2020 could have impacted the statewide burden of COVID-19 in Indiana. Specifically, we focused on the effects of school operating capacity and adherence to wearing face masks in schools. This focus was motivated by the fact that Indiana and other US states reopened their schools for in-person instruction in August with only minimal interventions of requiring face masks and physical distancing in schools, despite uncertainty about the proportion of students who would elect to attend in person and the degree to which they would adhere to face-mask and physical-distancing requirements. We approached this question with an agent-based model originally developed for pandemic influenza [30], which we tailored to SARS-CoV-2 [19, 31, 32, 33, 21, 34] and applied to a geographically and demographically realistic synthetic population representing Indiana. In addition to presenting outcomes across a range of hypothetical scenarios, we calibrated our model to data from the fall to assess the plausibility of K-12 school reopening as a driver of the observed resurgence of SARS-CoV-2 in Indiana during fall 2020.

## 2 Methods

### 2.1 Approach

Our approach to modeling SARS-CoV-2 transmission was based on the Framework for Reconstructing Epidemic Dynamics (FRED) [30], an agent-based model that offers the ability to explore the impacts of complex, non-pharmaceutical interventions in a natural way through modifications to individual behaviors. Using this model, we simulated the spread of SARS-CoV-2 in Indiana using a synthetic population with demographic and geographic characteristics of the state’s real population, including age, household composition, household location, and occupation [35]. We analyzed the impact of school reopening from August 24 (first day of classes in Marion County, the most populous) to December 31, 2020 in the population of Indiana as a whole, as well as in students, teachers, and their cohabitants. We quantified impact as the difference in the number of COVID-19 infections, symptomatic infections, and deaths between each scenario and a baseline scenario, the details of which differed according to which comparisons were of interest.

### 2.2 Agent-based model

Our model was based on a synthetic population of the entire state of Indiana [35], which included 1.3 million K-12 students, 1.7 million people living with students, and 6.3 million people in total (Table S10). Each of these agents visits a set of places defined by their activity space, which can include houses, schools, workplaces, long-term care facilities, and various neighborhood locations. Transmission can occur when an infected agent visits the same location as a susceptible agent on the same day, with numbers of contacts per agent specific to each location type. For example, school contacts depend not on the size of the school but on the age of the student and their assigned school grade, given that students have a higher number of contacts with students in their classroom than with those in other classrooms. Every day of the week, students and teachers visit their school, and students are assigned to classrooms based on their age. Given that schools are closed during the weekends, community contact is increased by 50% for students and teachers on weekends, unless they are sheltering in place [30]. Community contacts are modeled as contacts with other agents in the same neighborhood. For both schools and other locations, we adopted contact rates for each location type that were previously calibrated to influenza attack rates specific to each location type [30, 36].

Once infected, each agent had a latent period (mean = 3.35 days, standard deviation = 1.16 d) and an infectious period (mean = 3.7 d, s.d. = 1.2 d) drawn from distributions calibrated so that the average generation interval distribution matched estimates from Singapore (mean = 5.20 d, s.d. = 1.72 d) [33]. The absolute risk of transmission depended on the number and location of an infected agent’s contacts and a parameter that controls SARS-CoV-2 transmissibility upon contact, which we calibrated. A proportion of the infections were asymptomatic, with the probability of symptoms increasing with age [37, 19, 38] (Table S9, Fig. S4). We assumed that these infections were as infectious as symptomatic infections and had identical incubation and infectious period distributions [39, 40, 26, 41]. Furthermore, we assumed that children were less susceptible to infection than adults, which we modeled with a logistic function calibrated to model-based estimates of this relationship by Davies et al. [19]. We assumed that severity of disease increased with age, consistent with statistical analyses described elsewhere [32, 31, 38, 21].

Agent behavior in FRED has the potential to change over the course of an epidemic. Following the onset of symptoms, infected agents self-isolate at home according to a fixed daily probability, whereas others continue their daily activities [42, 43]. This probability was chosen so that, on average, 68% of agents will self-isolate at some point during their symptoms. This figure of 68% was based on the proportion of symptomatic infections among healthcare workers in the USA that developed fever during the course of their infection [43], and assuming that those with fever are likely to self-isolate. Agents can also engage in a variety of non-pharmaceutical interventions, including school closure, sheltering in place, and a combination of mask-wearing and physical distancing. School closures occur on specific dates across the state [44], resulting in students limiting their activity space to household and neighborhood locations on those days. Within households, agents interacted with their cohabitants on a daily basis. We assumed that agents did not wear face masks inside their homes, nor did they isolate from their household members if infected. To capture temporal changes in overall mobility and community contact over the course of the epidemic, we modeled a time-varying probability of sheltering in place. On days when an agent sheltered in place, they reduced their activity spaces to their home only, whereas other agents continued with their normal routines.

We modeled protection from face masks by reducing the probability of transmission when an infected agent wore a face mask. Our default assumption followed a median estimate of an adjusted odds ratio of 0.3 against SARS-CoV in non-healthcare settings [45, 34]. A meta-analysis by Chu et al. [34] included studies in healthcare settings in addition to those from non-healthcare settings, but we included only estimates that referred to the latter. The proportion of agents wearing face masks in workplace and community settings changed over time, and we assumed that agents did not wear face masks inside their households. In addition, students and teachers wore face masks according to probabilities specified as part of school reopening scenarios in our analysis. Further details about the model are available in the Supplementary Text.

### 2.3 Model calibration and validation

Two of the time-varying drivers of transmission in our model were informed by time-varying data inputs. First, we informed the daily probability of sheltering in place with mobility reports from Google [46]. In doing so, this sheltering-in-place probability in our model accounts for both the effects of shelter-in-place orders and some people deciding to continue staying at home after those orders are lifted [47]. Second, we informed the daily proportion of agents wearing face masks in workplace and community settings with Google Trends data for Indiana using the terms “face mask” and “social distancing” [48]. To inform the magnitude of the proportion of people wearing face masks in workplace and community settings, we used survey data [49] on face-mask usage from a single point in time.

The values of nine model parameters (listed in Table S9) were informed by calibrating the model to three time-varying epidemiological data streams corresponding to the state of Indiana— daily deaths, daily hospitalizations, and daily test positivity at the state level through August 10—and to the age distribution of cumulative deaths through July 13. We obtained daily incidence of reported cases and deaths from the New York Times COVID-19 database [1]. Daily hospitalizations and the age distribution of cumulative deaths were obtained from the Indiana COVID-19 dashboard [8]. Daily numbers of tests performed in the state were available from The Covid Project [50]. To calibrate the model to these data, we first used a Sobol design sampling algorithm [51, 52] to draw 6,000 combinations of the nine calibrated parameters. We then calculated the likelihood of each parameter combination given the data, and resampled the parameter combinations proportional to their likelihoods to obtain an approximation of the posterior distribution of parameter values. Additional details about the calibration procedure are described in the Supplementary Material.

To validate the model, we compared its predictions to data withheld from model calibration. Specifically, we compared the calibrated model’s predictions of the infection attack rate, both overall and by age, to results from two statewide serological surveys undertaken in late April and early June [53, 54]. We assessed the model’s success in this validation exercise by visual comparison of the model’s predictions with the data, with a focus on overlap between the 95% prediction intervals of the model and the 95% credible intervals associated with the empirically-derived estimates from the serological surveys.

### 2.4 Model outputs

The main outputs from our model were the numbers of infections, symptomatic cases, hospitalizations, and deaths at the state level. For different comparisons, these outputs were examined either on a daily basis, cumulatively between August 24 and December 31, or stratified by place of infection (school, home, other) or affiliation with schools (student, teacher, none). Another output that we examined was the daily reproduction number, *R*(*t*), which was calculated in the model for each infected agent and averaged across the modeled population. We defined *R*(*t*) as the average number of secondary infections caused by any agent who was infected on day *t*— i.e., the case reproduction number a la Fraser [55]. Finally, to account for uncertainty in these outcomes due to uncertainty in the calibrated parameters, we sampled parameter sets from our approximation of the posterior distribution of parameter values.

### 2.5 Model scenarios

#### 2.5.1 Effects of conditions in schools

To explore how alternative conditions for school reopening could have impacted the statewide burden of COVID-19 in Indiana, we performed simulations that spanned a range of assumptions about school operating capacity and adherence of students and teachers to face masks while in school. We chose to focus on these parameters given that they were two of the major unknowns as the state proceeded with its plans for in-person instruction beginning in August 2020. Given that students were offered the option of either in-person or remote instruction, we evaluated scenarios in which school operating capacity was either 50%, 75%, or 100%. Specifically, this parameter represents the daily probability that a student would go to school, such that all students could go to school at some point during the simulation but the average number of students in attendance on a given day is determined by the school operating capacity parameter. For each of these scenarios about school operating capacity, we also considered scenarios in which the adherence of students and teachers to face masks was either 50%, 75%, or 100%. In addition, we considered a scenario in which schools reopened normally (100% capacity, 0% face-mask adherence) and a scenario in which schools operated remotely (0% capacity, face-mask adherence irrelevant since no one in school).

#### 2.5.2 Sensitivity analyses

To explore the sensitivity of our results to model uncertainties, we performed three sets of sensitivity analysis. First, for each of the nine scenarios comprising our primary analysis, we analyzed the sensitivity of cumulative infections and the proportion of infections acquired in schools to each of the nine calibrated parameters by calculating partial rank correlation coefficients [56]. Second, we considered the sensitivity of our results to values of two assumed parameters not included in the calibration: protection afforded by face masks and the probability of isolation given symptoms. This included a total of four scenarios exploring lower and higher values of each of those parameters. Third, we considered alternative scenarios about select model assumptions that we regarded as potentially important unknowns about transmission of SARS-CoV-2 by children. These included scenarios in which asymptomatic infections (which are more likely to occur among children) are half as infectious as symptomatic infections, a scenario in which children aged 0-10 years have lower susceptibility (0.1), and a scenario in which individuals of all ages are equally susceptible to SARS-CoV-2 infection. For each of the alternative scenarios in the second and third sets of sensitivity analyses, we re-calibrated the model under that scenario and simulated it forward for the fall semester under the nine primary scenarios about school operating capacity and adherence to face-masks in schools. These latter two sets of sensitivity analyses all focused on cumulative infections statewide between August 24 and December 31, 2020.

#### 2.5.3 Retrospective analysis

To understand what conditions for school reopening resulted in model behavior consistent with data from fall 2020, we calibrated parameters for school operating capacity and face-mask adherence in schools to data from this time period. To do so, we held all other calibrated parameters at the values we estimated based on calibration to data from January 1 to August 24, 2020. Whereas we used Google mobility reports to drive a time-varying probability of sheltering in place during that initial period, we opted to assume fixed levels of mobility from August 24 through December 31. The main reason for this choice was that the Google data we used for the initial period showed a decrease in mobility during the fall, despite an increase in incidence, making it difficult to explain the epidemiological data under that assumption. To overcome that problem and to make this analysis more directly comparable to our other analyses, we held the probability of sheltering in place fixed at either of two levels of mobility during the fall: 1) mobility remained at summer levels, or 2) mobility increased to pre-pandemic levels. Under these assumptions, we calibrated the two focal parameters about conditions in schools to data from the fall using the same calibration procedure as we used during the initial period.

## 3 Results

### 3.1 Model calibration and validation

Our model was generally consistent with the data to which it was calibrated, capturing trends over time in daily deaths, hospitalizations, and test positivity at the state level (Fig. 1A-C), as well as greater proportions of deaths among older age groups (Fig. 1D). Some trade-offs in the model’s ability to recreate different data types were apparent, such as a recent increase in hospitalizations that the model failed to capture (Fig.1C), likely due to the predominance of data on deaths in the likelihood. Similarly, the model underestimated the proportion of deaths in people older than 80 (Fig. 1B), indicating a possible underestimation of the contacts in this age group in the overall community or in long-term care facilities. Even so, the model’s predictions reproduced the range of variability in the data, as assessed by the coverage probabilities of its 95% posterior predictive intervals (daily deaths: 0.85; daily hospitalizations: 0.93; daily test positivity: 0.95; cumulative deaths by age: 1.0). The model was also consistent with data withheld from fitting. Across all ages, the model’s 95% posterior predictive intervals of the cumulative proportion infected through late April (median: 0.017; 95% CrI: 0.0045-0.051) and early June (median: 0.022; 95% CrI: 0.0058-0.069) spanned estimates from two state-wide serological surveys [53] (Fig. 2A). Our model’s predictions also overlapped with age-stratified estimates from those surveys (Fig. 2B), although it underpredicted infections among individuals aged 40-60 years.

**Figure 1.**
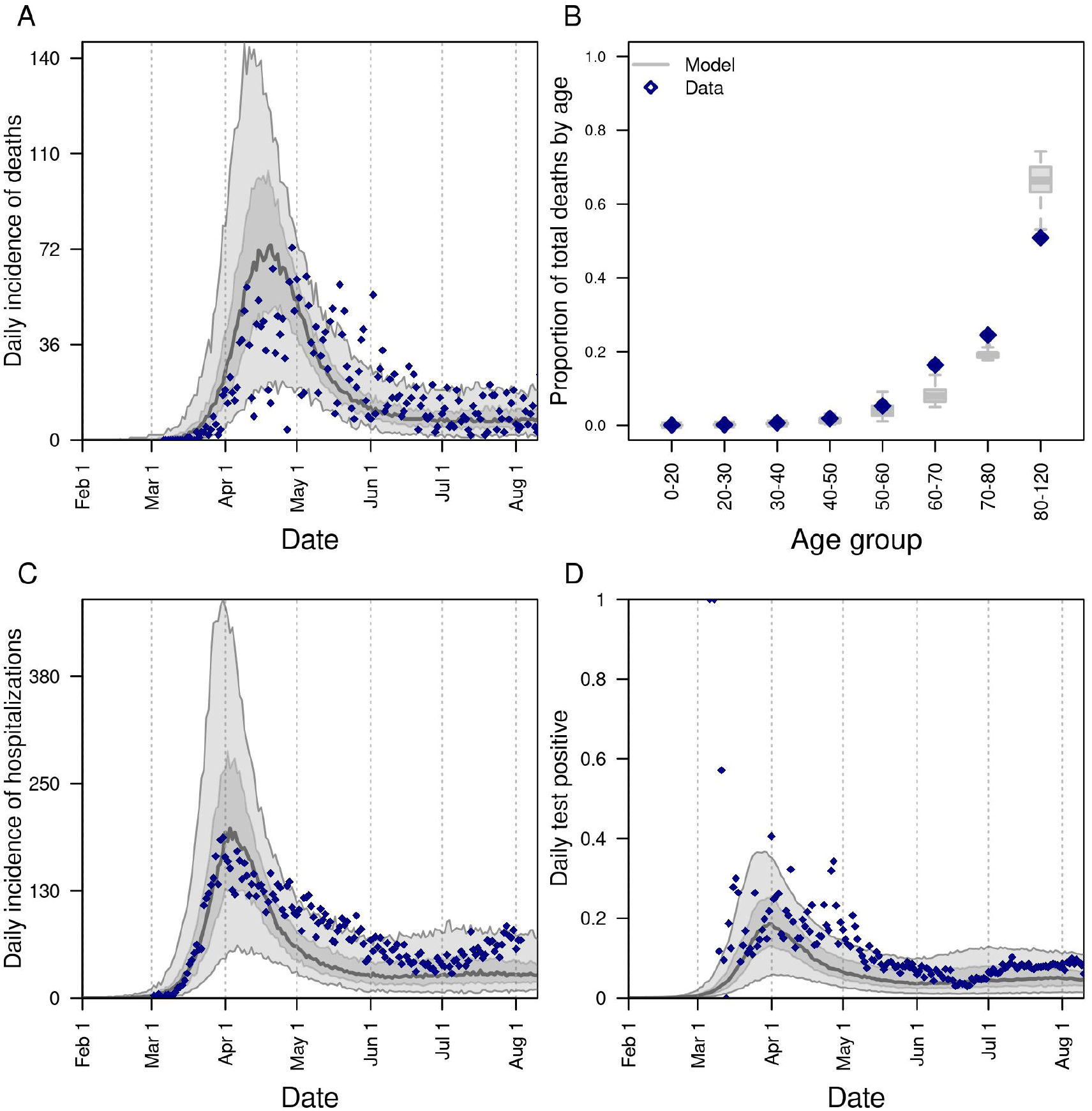
Model calibration to statewide data: A) daily incidence of death; B) proportion of deaths through July 13 in decadal age bins; C) daily incidence of hospitalization; and D) daily proportion of tests administered that are positive for SARS-CoV-2. In all panels, blue diamonds represent data. In A, C, and D, the gray line is the median, the dark shaded region the 50% posterior predictive interval, and the light shaded region the 95% posterior predictive interval.

**Figure 2.**
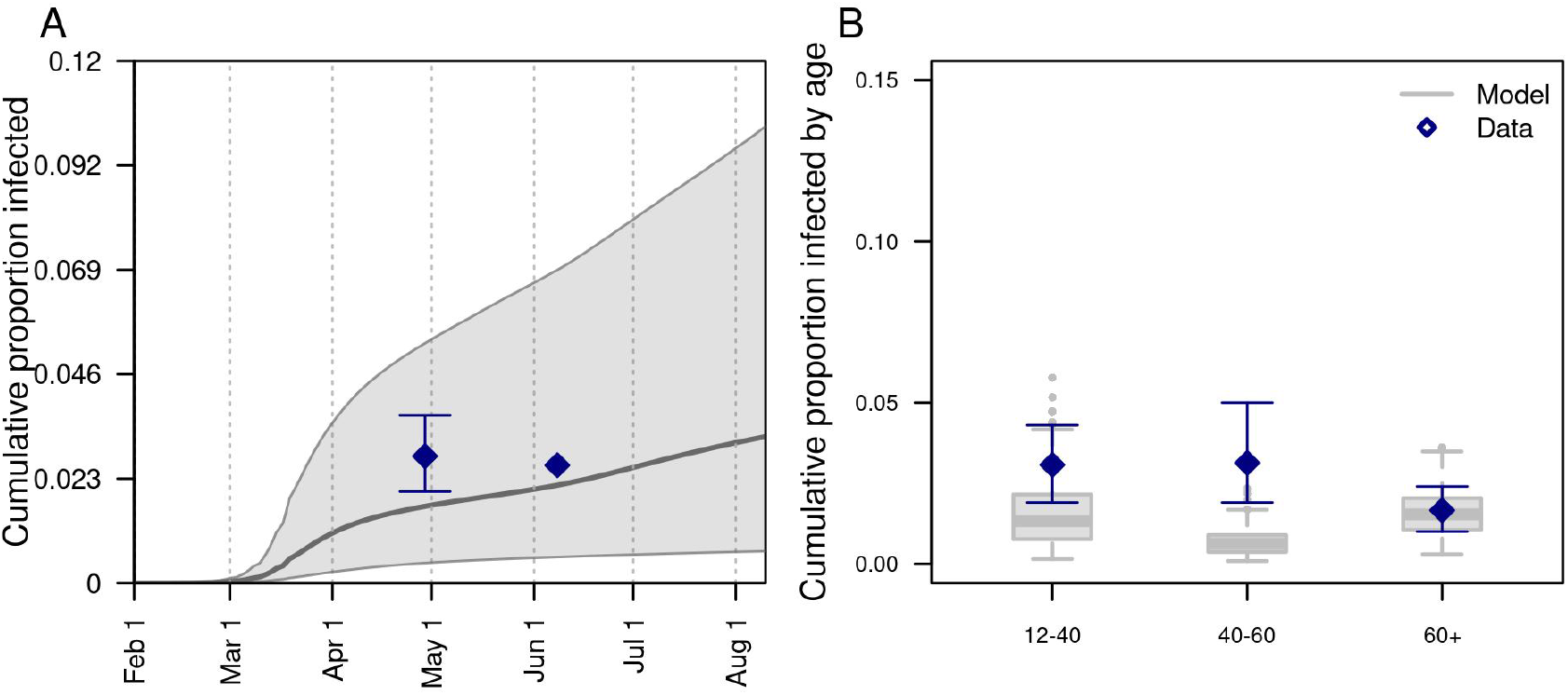
Model comparison with data withheld from fitting. We validated the model’s predictions against statewide data withheld from fitting on A) the cumulative proportion of the population of Indiana infected through late April and early June, and B) the cumulative proportion infected among individuals aged 12-40, 40-60, and 60+. Data are shown in navy and come from a random, statewide serological survey [53]. Model predictions are shown in gray. In A, the line and band indicate the median and 95% posterior predictive interval. In B, lines, boxes, and error bars indicate median, interquartile range, and 95% posterior predictive interval.

Calibration of the parameter that scaled the magnitude of SARS-CoV-2 importations [57, 58] in our model resulted in a median of 1.30 (95% PPI: 0.50-1.46) imported infections per day from February 1 to August 10. To ensure that the model reliably reproduced the high occurrence of deaths observed in long-term care facilities, we seeded infections into those facilities at a daily rate proportional to the prevalence of infection on that day; this calibrated proportion was 0.037 (95% PPI: 0.022-0.092). On the opposite end of the age spectrum, our calibration resulted in a median estimate of susceptibility among children of 0.346 (95% CrI: 0.311-0.506), compared to 0.834 (95% CrI: 0.652-0.946) in adults (Fig. S1). Our calibration resulted in an estimate of transmissibility (median: 0.593; 95% CrI: 0.501-0.788) that corresponded to values of *R*(*t*) during the initial phase of the epidemic in Indiana of 1.73 (95% CrI: 1.11-2.34), which represents an average of daily values across the first two weeks of March (Fig. 1A). This estimate is within the range of other estimates for *R*(*t*) that include the state of Indiana [59, 60]. Driven by a calibrated estimate that the proportion of people sheltering in place rose in early March and peaked at a median of 32.1% (95% CrI: 28.8-66.9%) on April 7 (Fig. S2A), our estimates of *R*(*t*) dropped to a low of 0.57 (95% CrI: 0.42-0.71) on April 7 and remained below 1 thereafter (Fig. 1A). Also impacting our estimates of *R*(*t*) was the increasing use of face masks in the community, which we estimated at 53.4% (95% CrI: 46.1-54.0%) as of July 19 (Fig. S2B). Note that this estimated distribution of community face-mask adherence does not differ between scenarios and is not the same as the level of adherence in schools, which we imposed at different levels depending on the scenario.

### 3.2 Effects of conditions in schools

#### 3.2.1 Effects on statewide burden

Under a scenario in which schools reopened at full capacity and without any use of face masks, our model projected that *R*(*t*) across the state as a whole would have increased to 1.72 (95% CrI: 1.43-2.17) by mid-September (Fig. 3A). Given our assumption that levels of sheltering in place and face-mask adherence in the community remained constant during the fall (Fig. S2B), this increase in transmission was driven by infections arising in schools (Fig. 3B). As a result, new infections statewide would have risen to levels in the fall far exceeding those from the spring (Fig. 3C). Under this scenario, our model projected a total of 2.57 million (95% CrI: 2.36-2.88 million) infections (Fig. 4A) and 10,246 (95% CrI: 7,862-13,794) deaths (Fig. 4B) from Indiana’s population as a whole between August 24 and December 31.

**Figure 3.**
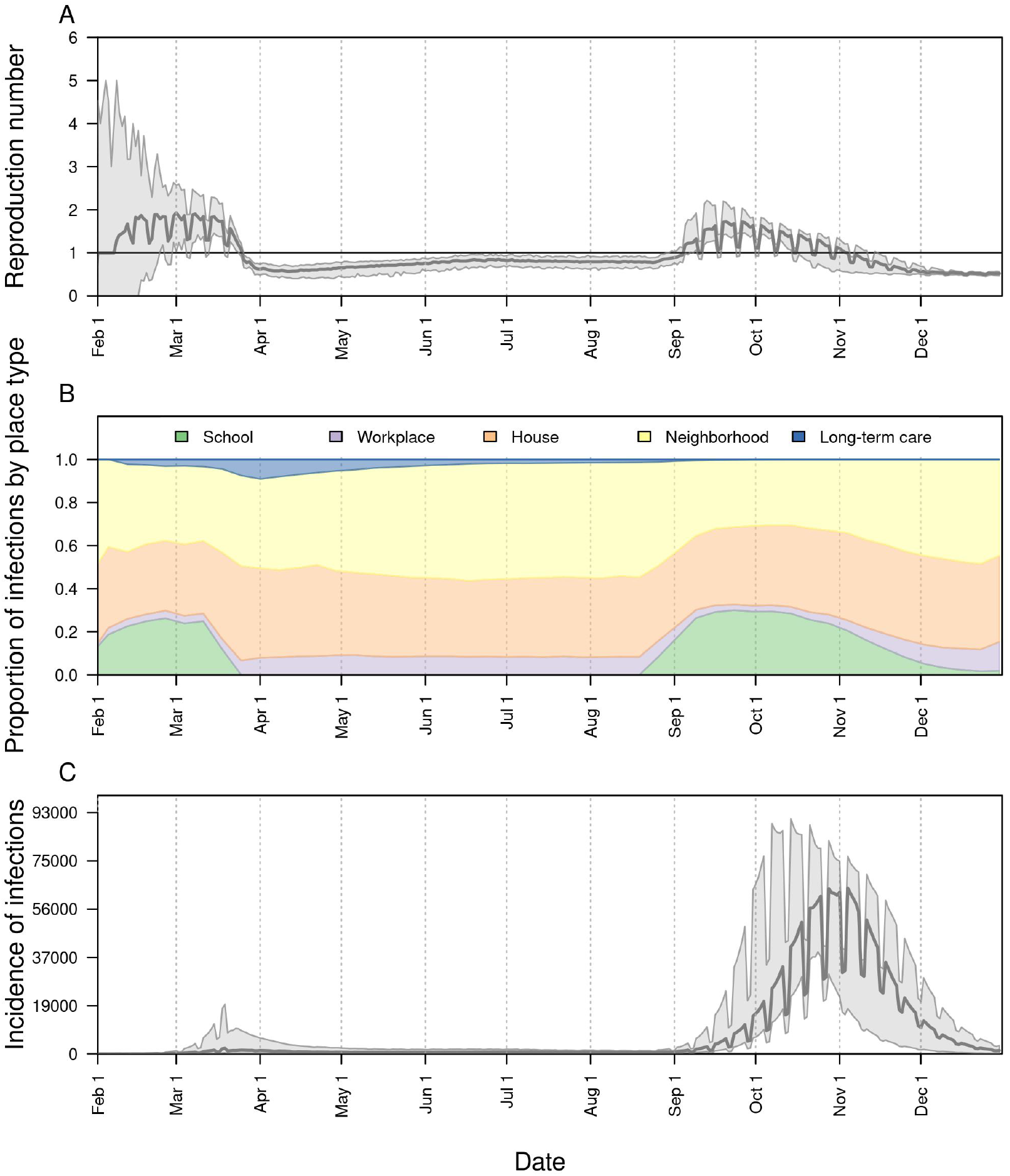
The impact of school reopening on August 24 under a scenario with 100% school operating capacity and 0% face-mask adherence in schools. Model outputs shown include: A) the reproduction number, *R*(*t*), over time; B) the proportion of infections acquired in different location types (colors) over time; and C) the daily incidence of infection statewide over time. In A and C, the line represents the median, and the shaded region represents the 50% posterior predictive interval.

**Figure 4.**
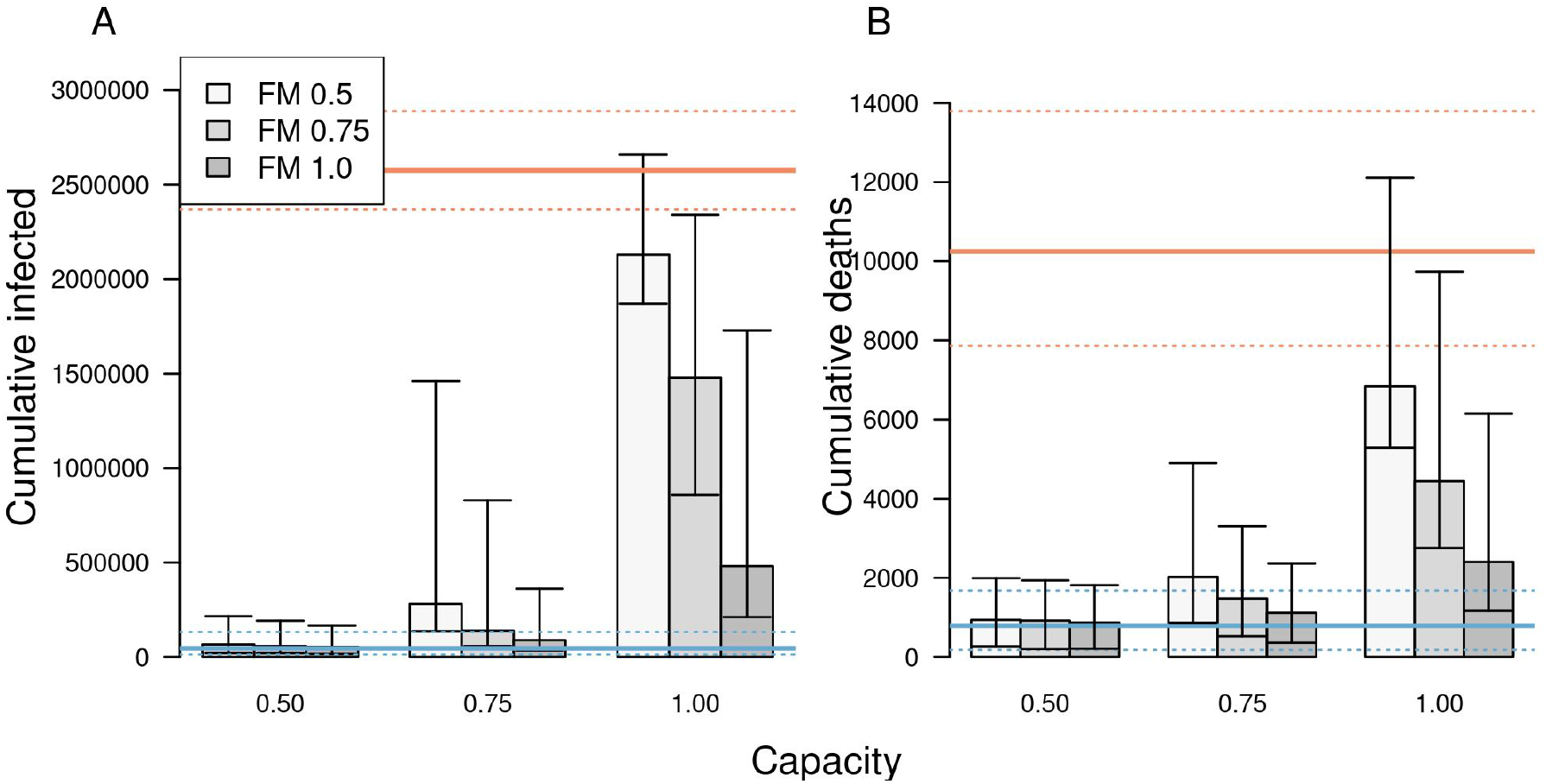
The impact of different scenarios about conditions for school reopening on A) cumulative infections and B) cumulative deaths in Indiana between August 24 and December 31. Scenarios are defined by school operating capacity (x-axis) and face-mask adherence in schools (shading). Orange lines represent projections under a scenario of school reopening at full capacity without masks (solid: median; dotted: 95% posterior predictive interval). Blue lines represent a scenario where schools operate remotely. Error bars indicate inter-quartile ranges.

Under a scenario in which schools went to remote instruction and all children remained at home, our model projected that *R*(*t*) would have remained near levels from August for the remainder of 2020 (Fig. S3A). This was a result of our assumption that sheltering in place and face-mask adherence in the community would have remained at their estimated levels as of August 13. Under this scenario, transmission would have continued through contacts at workplaces, within homes, and elsewhere in the community (Fig. S3B), resulting in a total of 45,579 (95% CrI: 14,651-132,546) infections (Fig. S3C) and 790 (95% CrI: 176-1,680) deaths from Indiana’s population as a whole between August 24 and December 31.

Less extreme scenarios about school operating capacity and face-mask adherence in schools also resulted in a wide range of variation in the projected statewide burden of COVID-19 during fall 2020. Under a scenario in which schools operated at 50% capacity and achieved 100% face-mask adherence, the cumulative numbers of infections and deaths that our model projected were similar to projections under a scenario in which schools operated remotely (Fig. 4, Tables S1 & S5). In general, cumulative infections and deaths statewide in fall 2020 were more sensitive to school operating capacity than to face-mask adherence in schools, with the worst outcomes projected to occur under a scenario with 100% school operating capacity and 50% face-mask adherence. Under this scenario, cumulative infections statewide were projected to have been 42.8 (95% CrI: 41.3-44.3) times greater than if schools had operated remotely, and cumulative deaths statewide were projected to have been 9.2 (95% CrI: 8.9-9.5) times greater (Table S1).

#### 3.2.2 Effects on risk for individuals affiliated with schools

Relative to a scenario with remote instruction, risk of infection and symptomatic infection was greatest for students (Figs. 5 & S5, left column), with a hundred-fold or greater increase in the risk of infection under a scenario with 100% school operating capacity and 50-75% face-mask adherence in schools (Tables S2 & S6). The risk of symptomatic disease in school-aged children was two-fold lower for children under 10 years of age (Fig. S6). Compared to students, the risk of infection was slightly lower for teachers, and much lower for students’ families. Due to their older ages, however, teachers and families experienced a much higher risk of death than students (Figs. 5 & S5, center & right columns). The highest risk of death was for teachers under scenarios with 100% school operating capacity (Figs. 5 & S5, center column). Compared to a scenario with remote instruction, the relative risk of death for teachers under these scenarios ranged from a 41-fold increase when face-mask adherence was 100% to a 166-fold increase when face-mask adherence was 50% (Table S3, S7). Under scenarios with 75% school operating capacity, those same relative risks dropped to a four-fold increase when face-mask adherence was 100% and a 22-fold increase when face-mask adherence was 50%. This again illustrates the overall greater effect of school operating capacity than face-mask adherence in schools.

**Figure 5.**
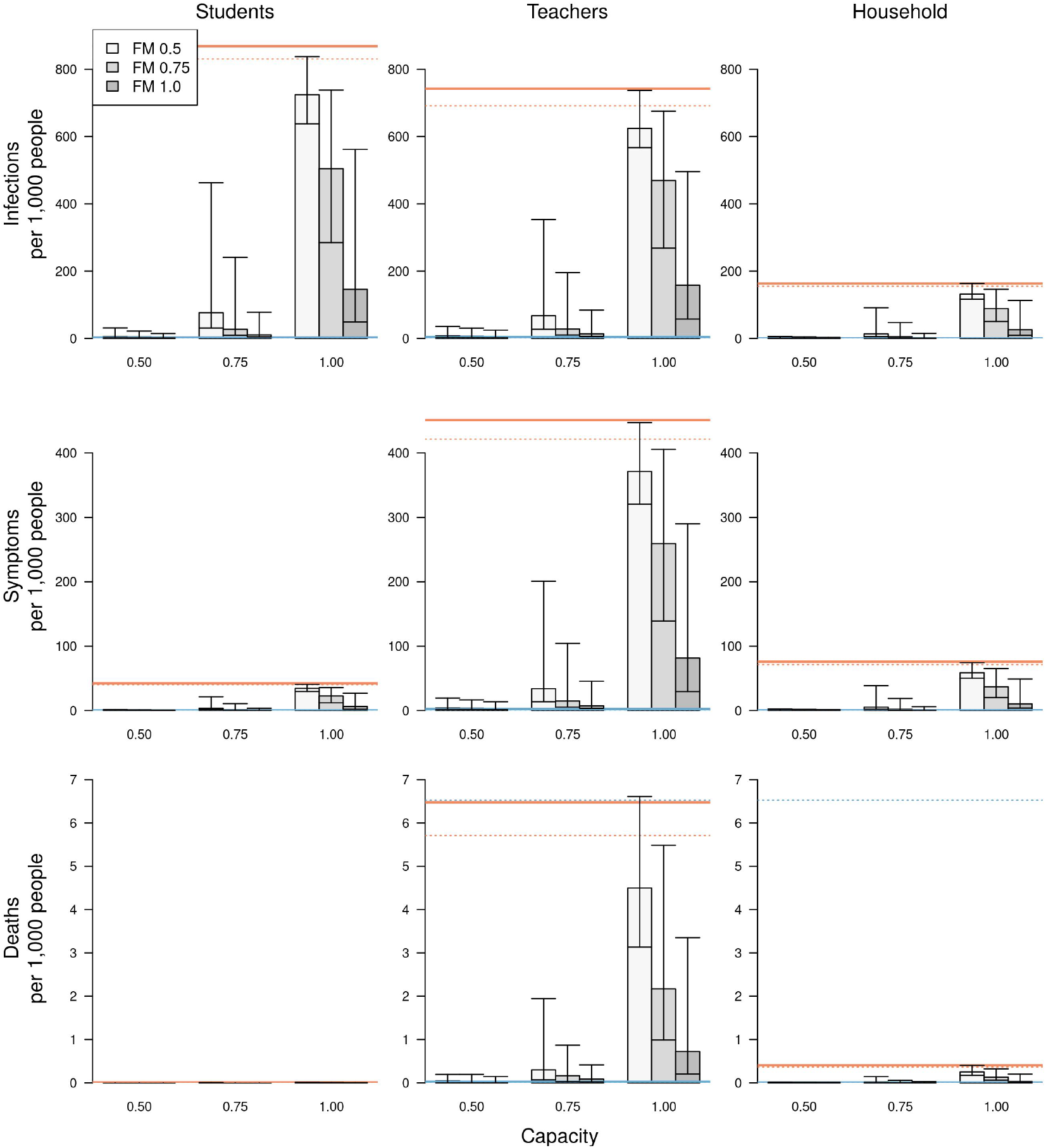
The impact of different scenarios about conditions for school reopening on infections (top row), symptomatic infections (middle row), and deaths (bottom row) per 1,000 people. These outcomes are presented separately for students (left column), teachers (middle column), and school-affiliated families (right column). Scenarios are defined by school operating capacity (x-axis) and face-mask adherence in schools (shading). Orange lines represent projections under a scenario of school reopening at full capacity without masks (solid: median; dotted: 95% posterior predictive interval). Blue lines represent a scenario where schools operate remotely. Error bars indicate inter-quartile ranges.

### 3.3 Sensitivity analyses

Our sensitivity analysis of the model’s nine calibrated parameters quantified the partial rank correlation coefficient (PRCC) of each of two model outputs: cumulative infections statewide from August 24 to December 31, and the proportion of infections acquired in schools. In general, these outputs were most sensitive to parameters controlling the age-susceptibility relationship (Figs. S7 & S8). Under some scenarios, the minimum susceptibility parameter, which applied to young children, had a PRCC as high as 0.6. The transmissibility parameter also had a PRCC that high, but only in some scenarios. For example, in scenarios with lower school operating capacity, the two parameters most relevant to community transmission—transmissibility and face-mask adherence in community settings—had a greater influence on cumulative infections statewide (Fig. S7), given that the contribution of schools to transmission was diminished in those scenarios.

Our sensitivity analysis of parameters for protection afforded by face masks and the probability of isolation given symptoms focused on cumulative infections statewide in fall 2020. For each alternative value of these parameters, we re-calibrated the model and generated projections under our nine primary scenarios about school operating capacity and face-mask adherence in schools. In general, the relative effects of differences in face-mask adherence in schools and school operating capacity were similar under alternative values of protection afforded by face masks (Table S11). However, relative to a baseline with schools operating remotely, the magnitude of the proportional increase in cumulative infections was sensitive to the level of protection afforded by face masks. For example, under a scenario with 75% face-mask adherence and 75% school operating capacity, the increase in cumulative infections ranged from 23.5-fold to 1.6-fold across the range of values of protection afforded by face masks that we explored (adjusted odds ratio = 0.12-0.73). Proportional increases in cumulative infections were generally insensitive to the probability of isolation given symptoms (Table S12). For example, under a scenario with 75% face-mask adherence and 75% school operating capacity, the increase in cumulative infections ranged from 4.9-fold to 6.9-fold across the range of values of isolation probability that we explored (0.5-0.9).

Our sensitivity analysis of assumptions related to the role of children in transmission also focused on cumulative infections statewide in fall 2020. Relative to a baseline with schools operating remotely, the proportional increase in cumulative infections was very similar to our default assumptions under a scenario with lower susceptibility among children aged 0-10 years and a scenario with lower infectiousness of asymptomatic infections (Table S13). This was the case for all scenarios about face-mask adherence in schools and school operating capacity, except for the most extreme case in which face-mask adherence in schools was 50% and school operating capacity was 100%. In that case, our default assumptions resulted in a 42.9-fold increase in cumulative infections, whereas the two alternative scenarios resulted in a 26-fold increase. Under a scenario with equal susceptibility for all ages, proportional increases in cumulative infections were much higher than under our default assumptions (Table S13). Because susceptibility in children was higher under this scenario, transmission statewide was much higher when school reopened, especially in scenarios with higher school operating capacity (Fig. S9). By the same token, prevalence dropped to levels over the summer when school was not in session that were much lower than the data suggest (Fig. S10), which raises doubts about the plausibility of this scenario.

### 3.4 Retrospective analysis

The model successfully reproduced statewide data from fall 2020 under relatively high values of school operating capacity and intermediate values of face-mask adherence in schools (Fig. 6). Under an assumption that the probability of sheltering in place in the state as a whole was fixed at levels from summer 2020 (Fig. 6, gray), the model calibration resulted in a median school operating capacity of 0.88 (95% CrI: 0.8-0.95) and a median face-mask adherence in schools of 0.6 (95% CrI: 0.4-0.7). Under an assumption that the probability of sheltering in place was fixed at pre-pandemic levels (Fig. 6, red), the model calibration resulted in a median school operating capacity of 0.85 (95% CrI: 0.77-0.92) and a median face-mask adherence in schools of 0.61 (95% CrI: 0.41-0.75). Overall, there was a wide range of values of face-mask adherence in schools that were consistent with the data, and the range of values of school operating capacity consistent with the data depended somewhat on face-mask adherence in schools (Fig. 6, bottom left).

**Figure 6.**
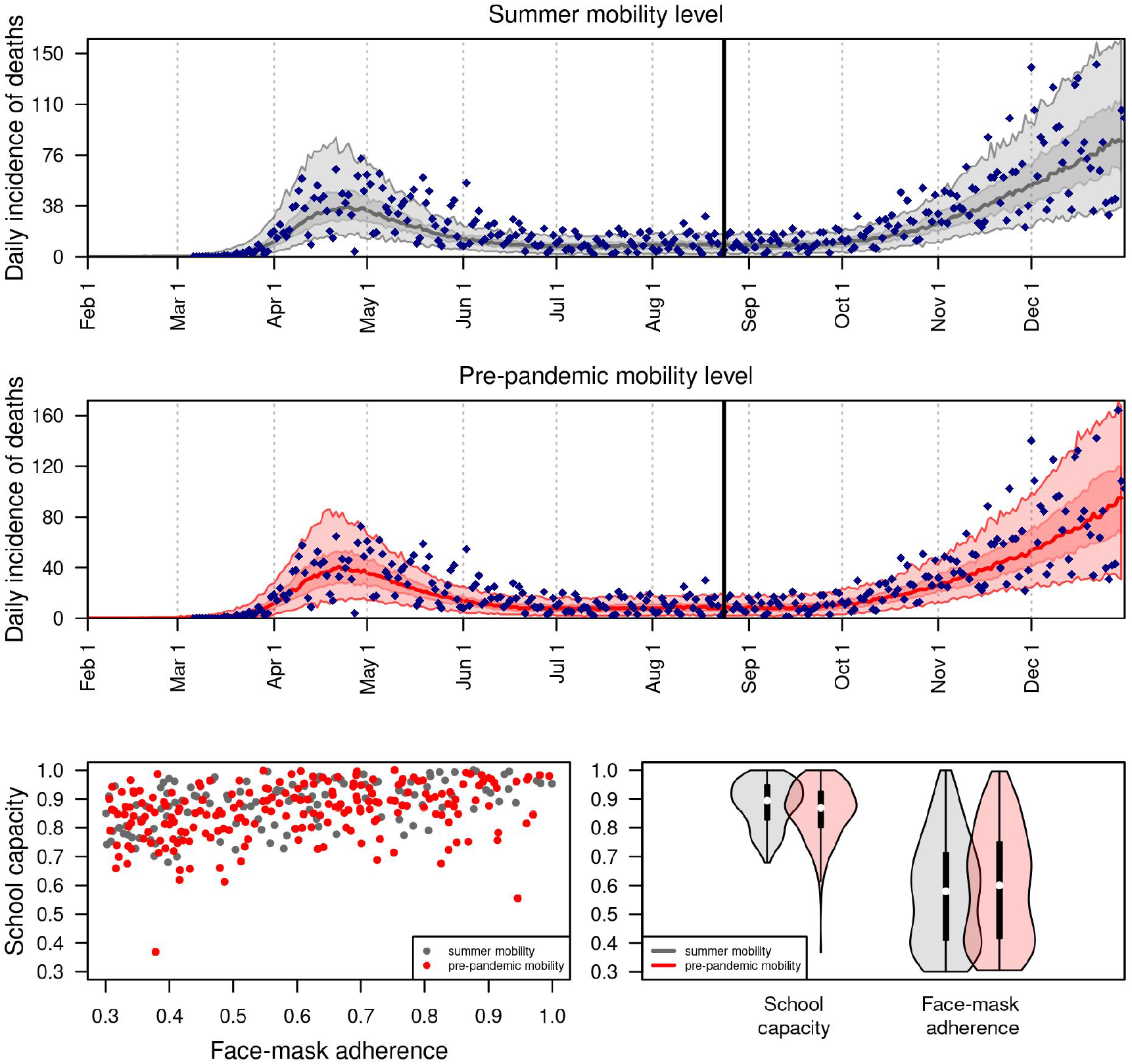
Retrospective analysis of the model calibrated to statewide data from fall 2020. Under two alternative scenarios about the daily probability of sheltering in place (summer vs. pre-pandemic mobility level in gray and red, respectively), we calibrated the parameters for school operating capacity and face-mask adherence in schools to data from August 24 through December 31, 2020 (blue diamonds). The calibrated model’s correspondence to daily incidence of death statewide is shown in the top two panels, and values of the calibrated parameters are shown in the bottom panels.

## 4 Discussion

Our model provides a detailed, demographically realistic representation of SARS-CoV-2 transmission in Indiana that is consistent both with data to which it was calibrated and to data that was withheld from calibration. In contrast to models that rely on assumptions about intervention impacts or estimate them statistically [2, 61], our model makes predictions about intervention impacts based on first-principles assumptions about individual-level behavior and contact patterns. Consistent with results from other analyses [2, 61, 20], the inputs and assumptions in our model led to a prediction that schools made a considerable contribution to SARS-CoV-2 transmission in February and early March, prior to large-scale changes in behavior. Extending that, a primary result of our analysis is that K-12 school reopening was capable of making a considerable contribution to SARS-CoV-2 transmission during fall of 2020, with the degree of that contribution dependent on conditions in schools.

The burden of COVID-19 associated with in-person school operation was predicted by our model to fall unevenly across the state’s population. In scenarios of school operating capacity of 100% with 50% face-mask adherence in schools, our model predicted that hundreds of thousands of children could have been infected during the fall semester, with very few of those resulting in deaths. In contrast, our results show that hundreds of deaths in teachers and school-affiliated families could have occurred. Our model indicates that the burden of COVID-19 in schools, teachers, and school-affiliated families across the state could have been reduced by operating at reduced capacity and achieving high face-mask adherence in schools. Under the relatively optimistic scenario of 50% school operating capacity and 100% face-mask adherence in schools, our model predicted that infections and deaths statewide would have been only 22% greater than under a scenario with fully remote instruction. In contrast, our model results suggest that if schools would have operated at full capacity, infections and deaths statewide could have been one to two orders of magnitude greater than the scenario with fully remote instruction, especially with poor face-mask adherence in schools. When we extended our model calibration to account for data from fall 2020, we found that school operating capacity of 80-95% and face-mask adherence in schools of 40-70% resulted in model predictions most consistent with the observed data. For reference, data from the National COVID-19 School Response Dashboard [62] indicate that school operating capacity in Indiana was 77-83% during the fall. Although conditions elsewhere in the community likely played a role in statewide trends during the fall and were not accounted for fully by our model, these results demonstrate that transmission associated with K-12 school reopening was capable of driving the statewide resurgence of COVID-19 observed in Indiana in fall 2020.

The impacts associated with reduced school operating capacity result from reductions in both the number of contacts within the school and the probability that an infected student would be in attendance in the first place, similar to the logic behind why smaller gatherings are associated with reduced risk of transmission [3, 63, 64]. The magnitude of our results was most sensitive to the degree of protection afforded by face masks, which remains uncertain in school and other community settings for SARS-CoV-2 [34]. That uncertainty can be reduced as more studies are conducted. Recently, some studies have shown that face masks offer significant protection in community settings similar to what we assumed. For example, Payne et al. followed 382 U.S. Navy service members who reported wearing face masks and found a reduced risk of 70% in those who did [65]. Similar values were found in studies of 124 households in China [66] and 839 close contacts of 211 index cases in Thailand [67].

Although the scenarios we considered resulted in projected impacts spanning nearly the full range between fully remote instruction and fully in-person instruction with no face masks, they are a simplification of the complexities of how schools likely operated in fall 2020. Scenarios that we did not explore include different groups of students attending in person or remotely [68], varying degrees of modularization within schools [69], and the implementation of testing-based control strategies in schools [70]. In the event that infectiousness is lower for asymptomatic infections, the impact of school reopening on lower grades could have been lower than our results suggest. A related simplification of our statewide analysis is that the state, in reality, consists of a patchwork of policies across districts. In light of this complexity that our model does not capture, our results should be interpreted with caution in relation to specific counties or school districts below the state level. Across all scenarios though, our results illustrate the importance of reduced school operating capacity and maximal face-mask adherence in schools, as do other modeling studies [68, 69, 70, 71, 72].

A critical assumption of our analysis is that children are capable of being infected with SARS-CoV-2 and transmitting it to others at meaningful levels. Although the burden of severe disease skews strongly towards older ages [22, 73, 8], there are other lines of evidence that support our assumption. These include a contact-tracing study that found no distinguishable difference between infectivity of children and adults [26], several studies that found no distinguishable difference in viral load between children and adults [74, 40, 39, 75], a study that observed a greater secondary attack rate among children in homes [26], and a modeling study that found no evidence that children were less infectious [76]. More direct evidence comes from COVID-19 outbreaks that have been observed in schools, such as one in a high school in Israel in which 13.2% of students and 16.6% of staff were infected in just 10 days [77]. Even more pertinent, since schools reopened in September 2020, 31,658 COVID-19 cases have been reported in students and 13,240 cases have been reported in teachers and school staff across the state, as of April 2021 [8].

There is now a growing body of evidence that school closures contributed to mitigating the first wave of the epidemic [6, 70] and, as we have shown, may have contributed to the resurgence of SARS-CoV-2 during fall 2020. Our study adds to this evidence, and suggests an even greater impact of school reopening than several other studies [72, 70, 69, 78, 68]. This is due in part to our assumption that asymptomatic and symptomatic infections contribute similarly to transmission [26, 74, 40, 39, 75], and in part to our model’s ability to capture chains of transmission within schools and extending out into the community. Our study echoes several modeling studies in emphasizing the importance of reducing school operating capacity to impede transmission [69, 70, 71, 72, 78]. As schools grapple with COVID-19 going forward, results such as these provide an important basis for motivating the adoption and sustainment of reduced school operating capacity and adherence to face-mask requirements in schools. As we demonstrated, these actions are highly consequential for those directly linked to schools and for the communities in which they are embedded.

## Data Availability

The data is publicly available

## 5 Acknowledgements

This work was supported by an NSF RAPID grant to TAP (DEB 2027718), an Arthur J. Schmitt Fellowship and Eck Institute for Global Health Fellowship to RJO, and a Richard and Peggy Notabaert Premier Fellowship to MP. We thank the University of Notre Dame Center for Research Computing for computing resources.

## Supplementary material

**Figure S1.**
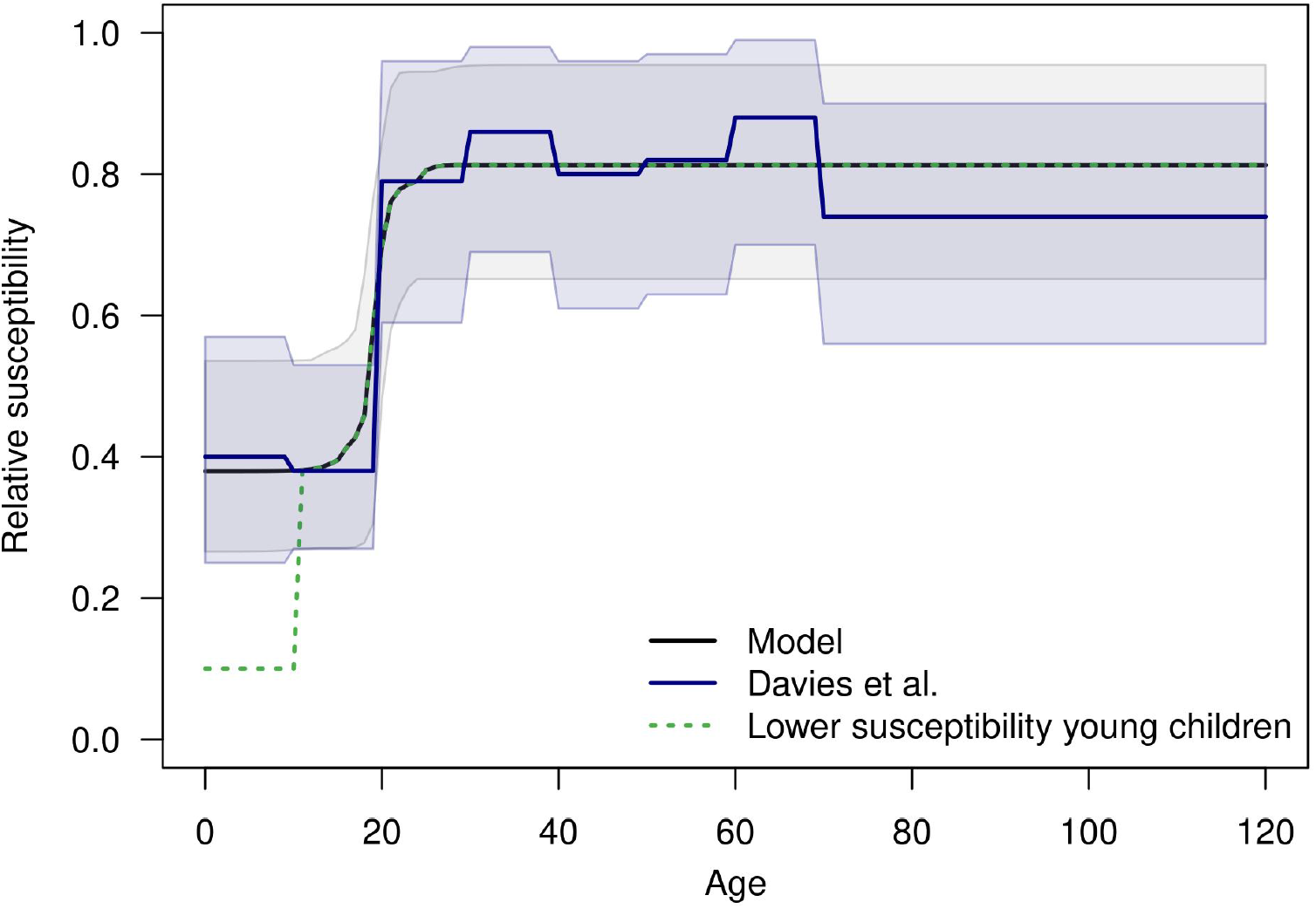
Susceptibility to SARS-CoV-2 infection by age. The black line shows the median of the estimated curve of susceptibility by age, which took the form of a modified logistic function defined by four parameters: minimum, maximum, inflection point, and slope. The gray band represents the 95% credible interval. Navy lines show estimates from Davies et al. [19] that were used to inform our estimates. The dashed green line shows an alternative scenario with lower susceptibility for children under 10 years of age.

**Figure S2.**
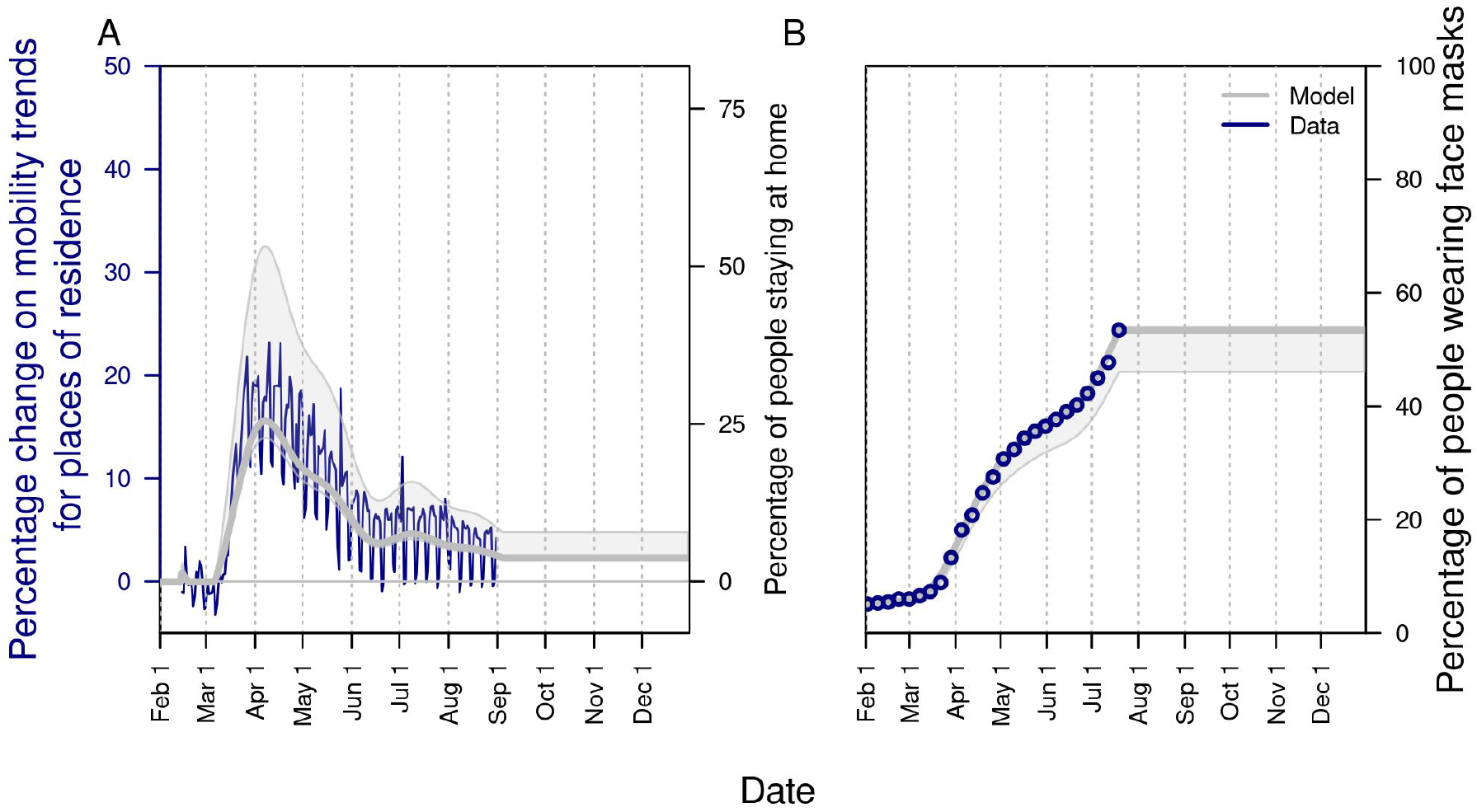
Changes in A) mobility patterns over time and B) face-mask adherence in the community as a whole. In A, the gray line shows the fitted pattern for the Google mobility index related to residential locations (navy line). Adherence to shelter-in-place in the model follows the same trend, but with its magnitude estimated through the model calibration process. In B, the gray line is informed by fitting to Google search data on “face mask” and assuming that values plateau from July 19 onward at a value informed by survey data [48, 49].

**Figure S3.**
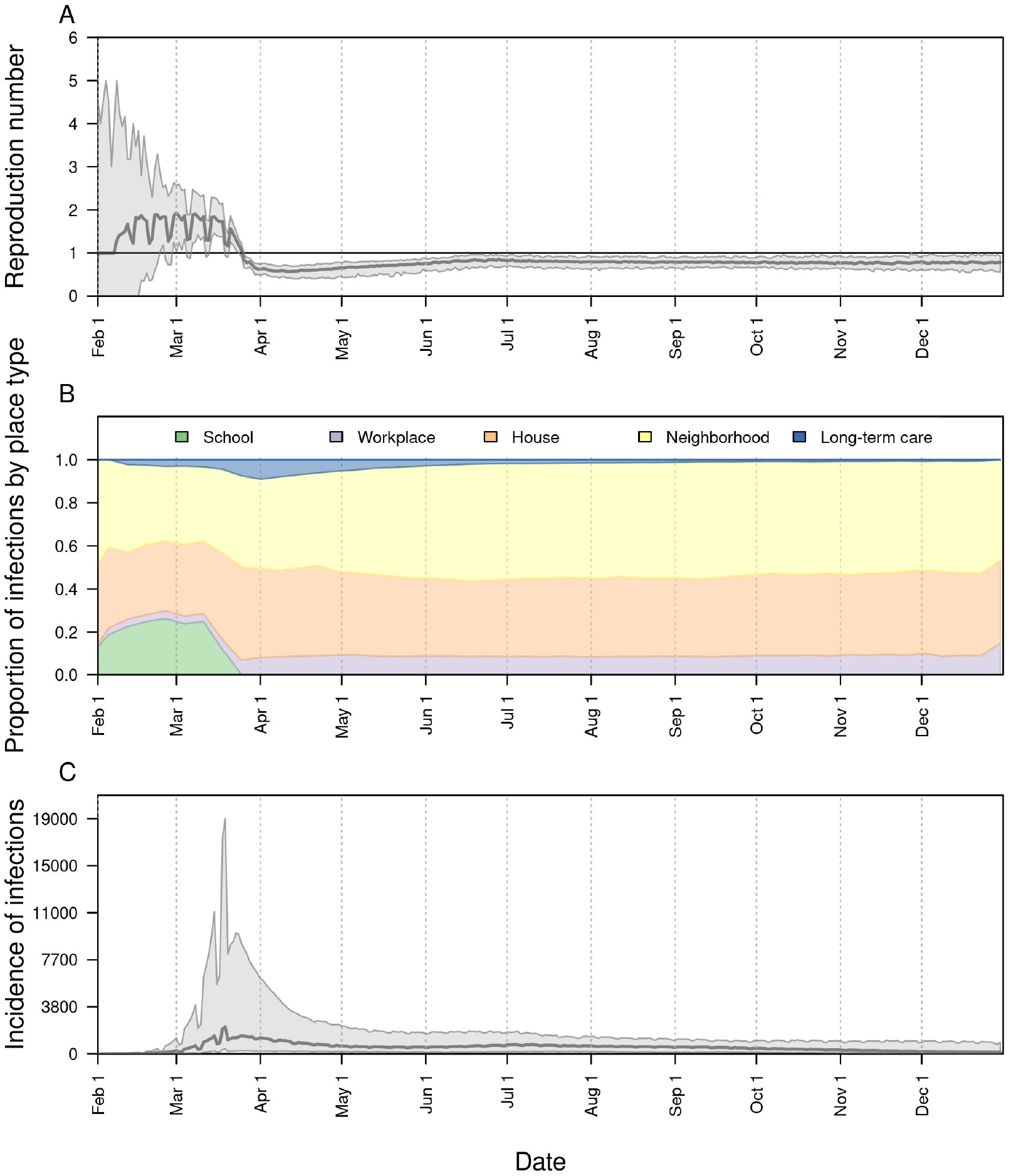
The impact of remote instruction (i.e., 0% school operating capacity). Model outputs shown include: A) the reproduction number, *R*(*t*), over time; B) the proportion of infections acquired in different location types (colors) over time; and C) the daily incidence of infection statewide over time. In A and C, the line represents the median, and the shaded region represents the 50% posterior predictive interval.

**Figure S4.**
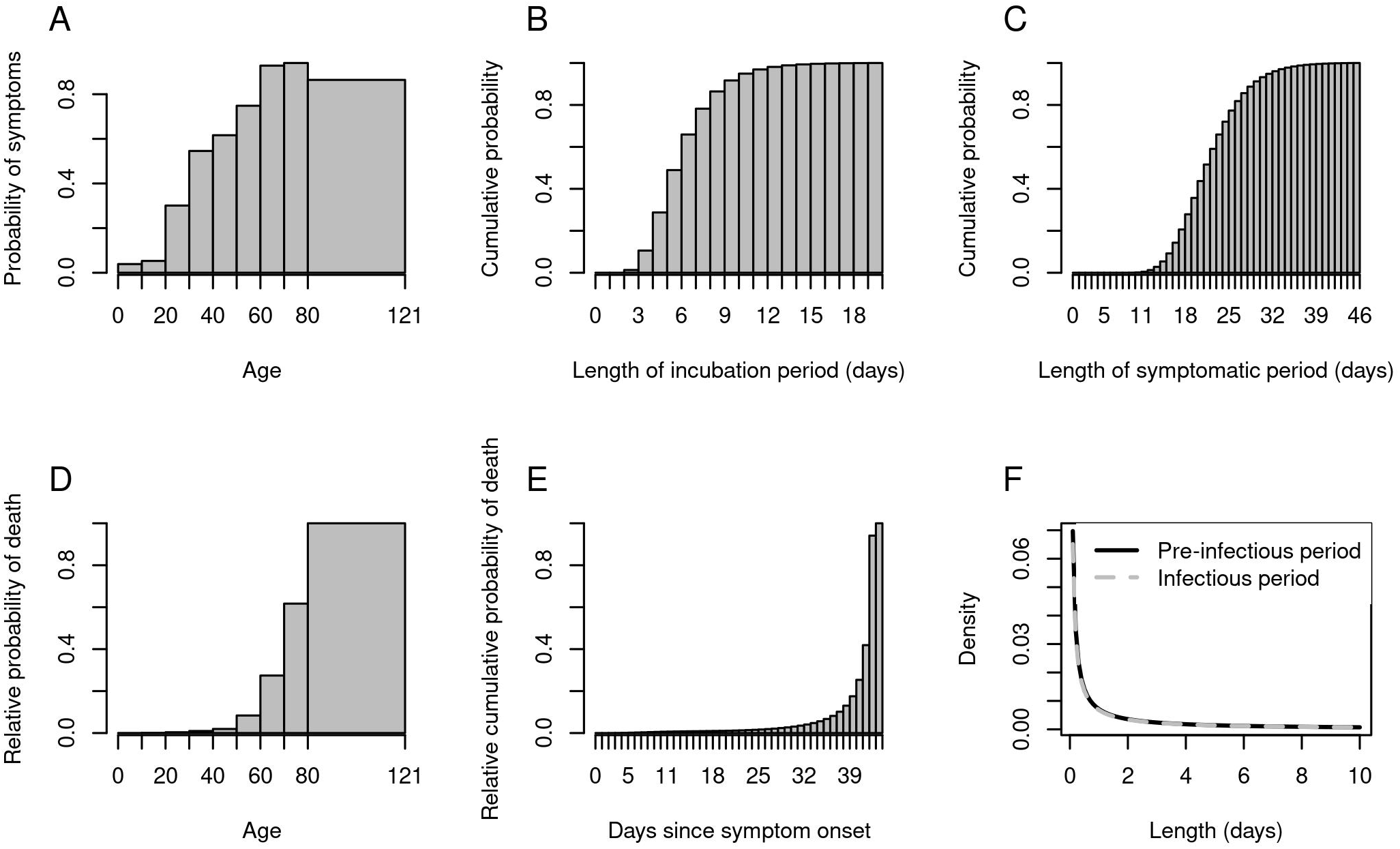
Distributional representations of select model parameters: A) probability of symptoms by age group; B) cumulative probability of the duration of the incubation period; C) cumulative probability of the duration of the symptomatic period; D) relative probability of death by age; E) cumulative probability of death by day after symptom onset; and F) density of duration of the pre-infectious and infectious periods.

**Figure S5.**
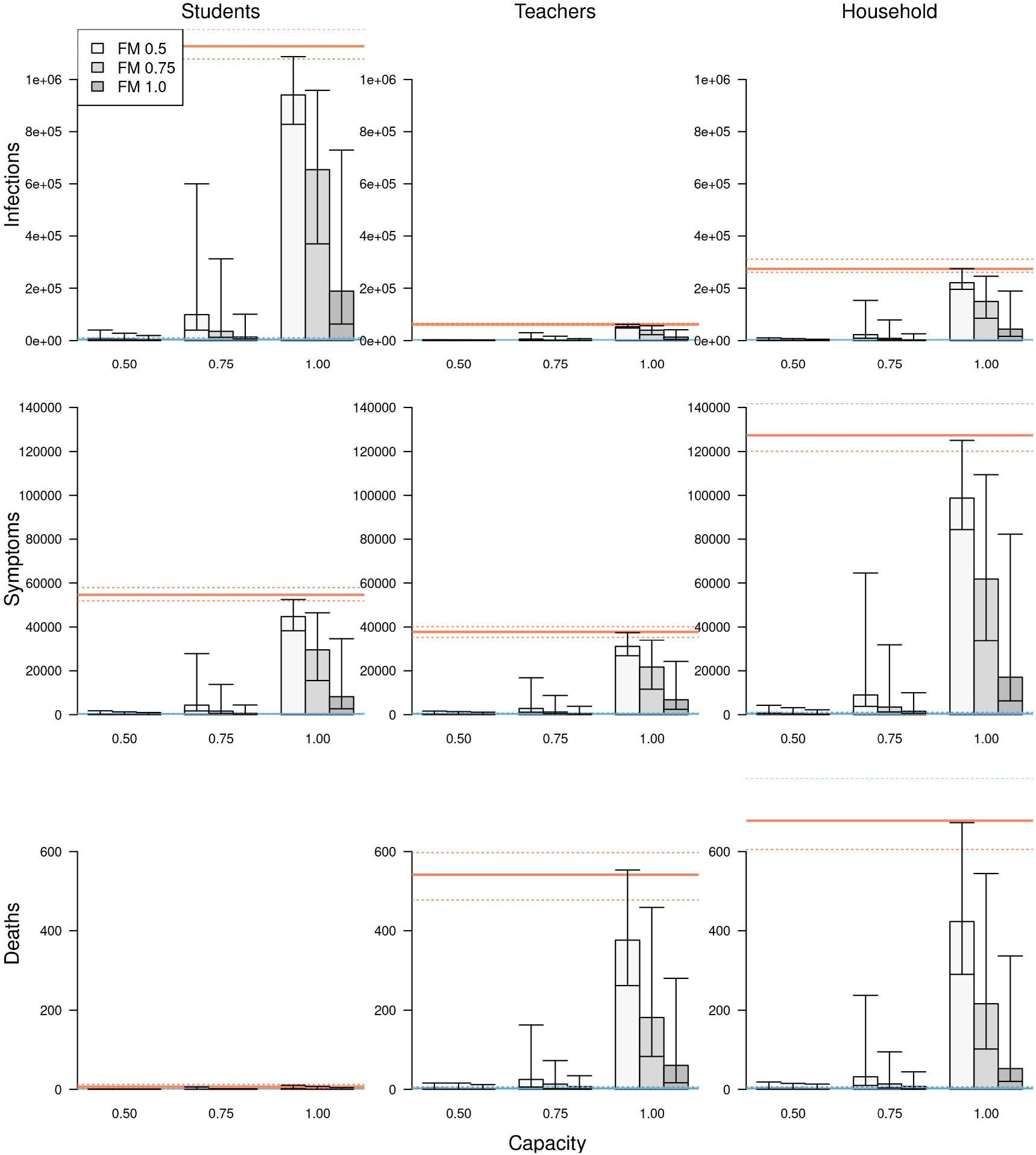
The impact of different scenarios about conditions for school reopening on infections (top row), symptomatic infections (middle row), and deaths (bottom row) cumulatively between August 24 and December 31 across the state of Indiana. These outcomes are presented separately for students (left column), teachers (middle column), and school-affiliated families (right column). Scenarios are defined by school operating capacity (x-axis) and face-mask adherence in schools (shading). Orange lines represent projections under a scenario of school reopening at full capacity without masks (solid: median; dotted: 95% posterior predictive interval). Blue lines represent a scenario where schools operate remotely. Error bars indicate inter-quartile ranges.

**Figure S6.**
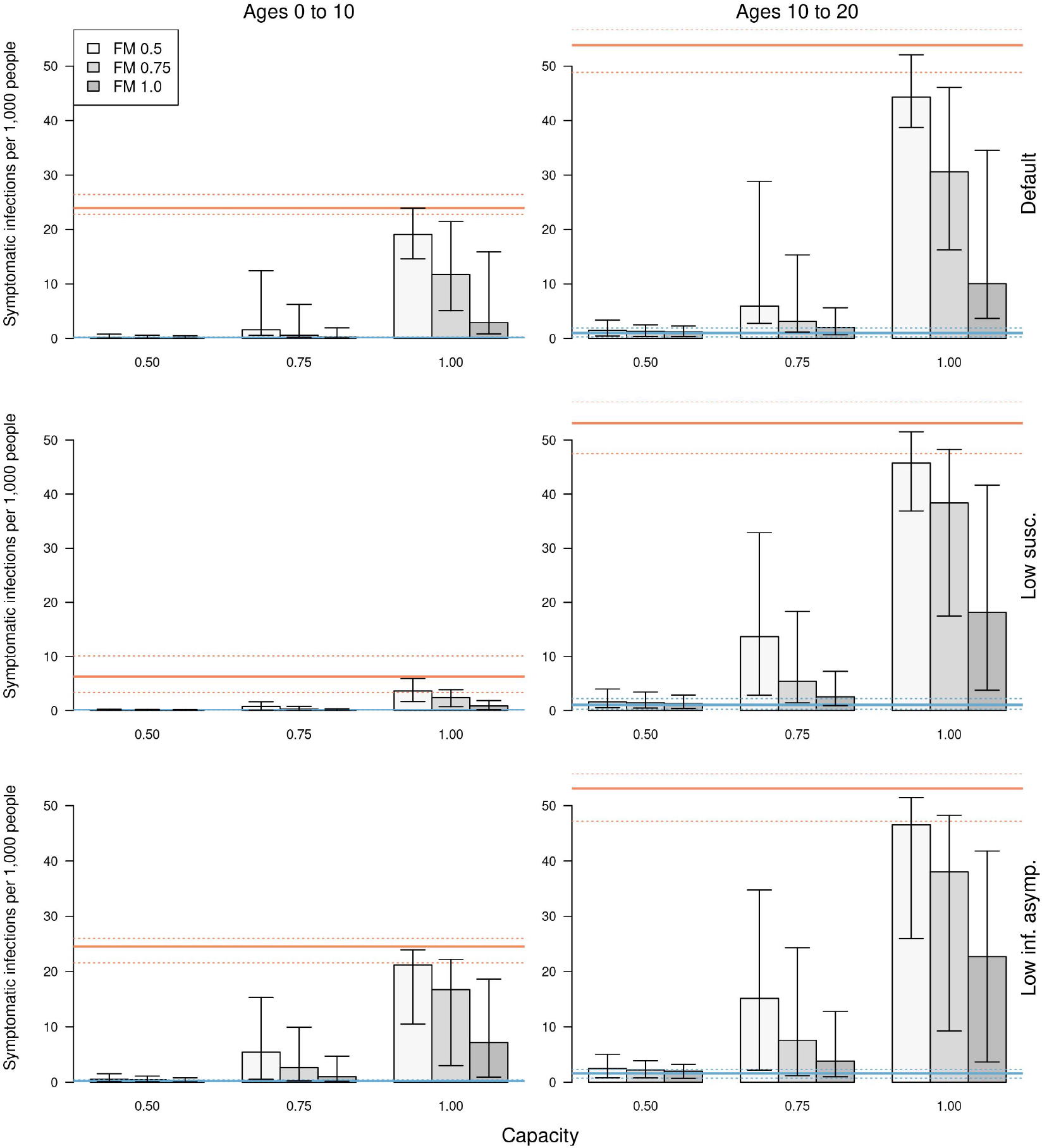
The impact of different scenarios about conditions for school reopening on cumulative symptomatic infections per 1,000 people for ages 0-10 (left column) and 10-20 (right column). Results are shown under: (top row) the baseline assumption for susceptibility and infectiousness of asymptomatic infections; (middle row) lower susceptibility for children under 10 years of age; and (bottom row) the baseline assumption for susceptibility but with lower infectiousness of asymptomatic infections. Scenarios are defined by school operating capacity (x-axis) and face-mask adherence in schools (shading). Orange lines represent projections under a scenario of school reopening at full capacity without masks (solid: median; dotted: 95% posterior predictive interval). Blue lines represent a scenario where schools operate remotely. Error bars indicate inter-quartile ranges.

**Figure S7.**
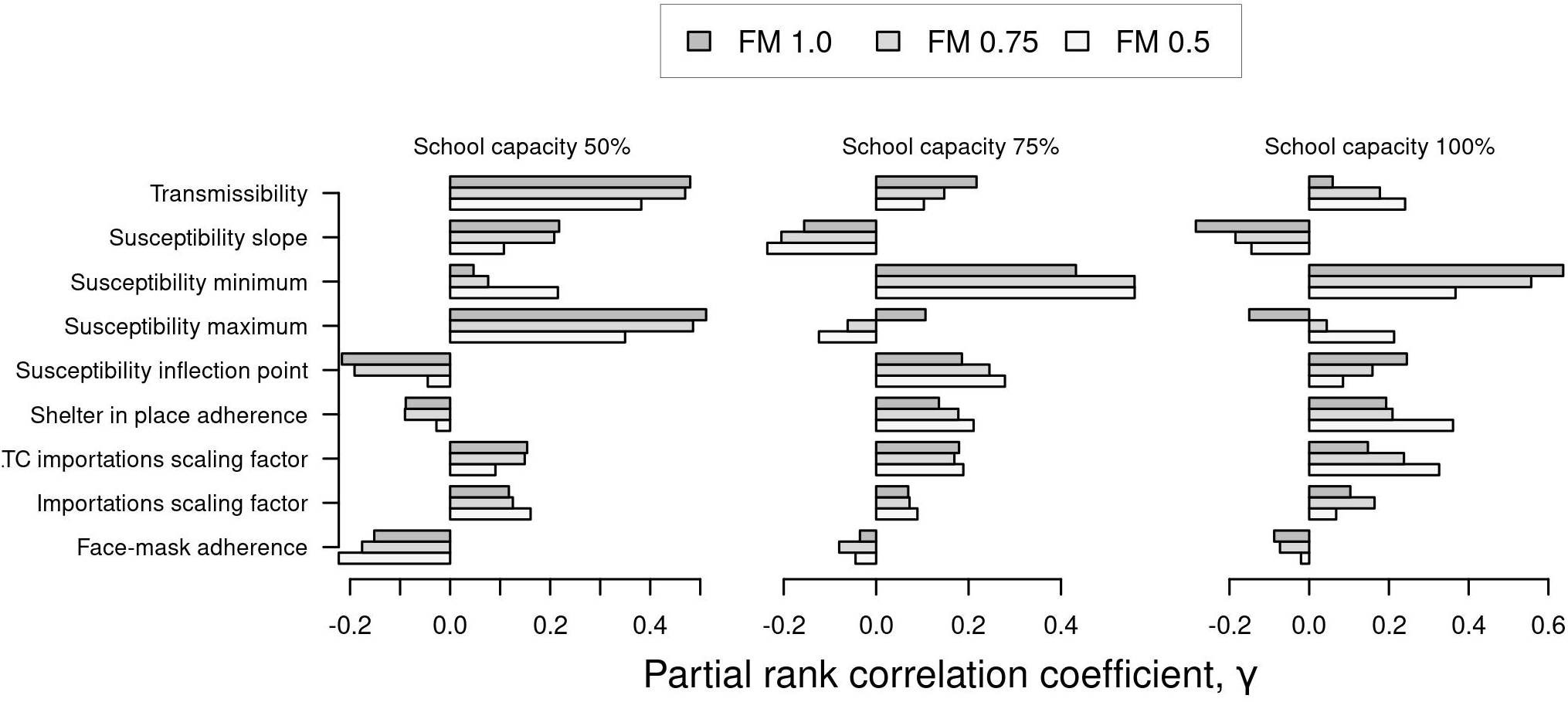
Sensitivity analysis of cumulative infections statewide between August 24 and December 31 to variation in the model’s nine calibrated parameters. Bars indicate values of the partial rank correlation coefficient. Results are presented separately by school operating capacity (panels) and face-mask adherence in schools (gray shading).

**Figure S8.**
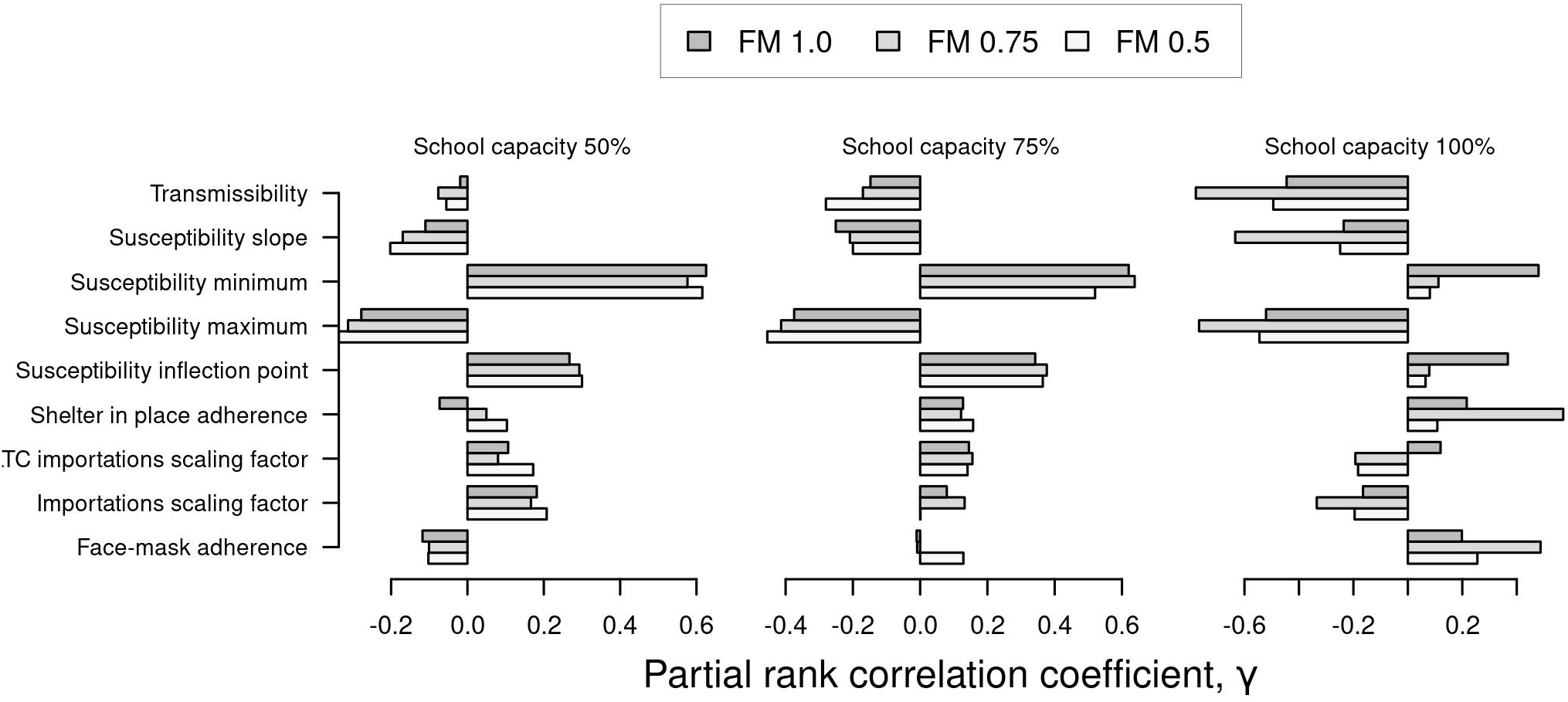
Sensitivity analysis of the proportion of infections acquired in schools between August 24 and December 31 to variation in the model’s nine calibrated parameters. Bars indicate values of the partial rank correlation coefficient. Results are presented separately by school operating capacity (panels) and face-mask adherence in schools (gray shading).

**Figure S9.**
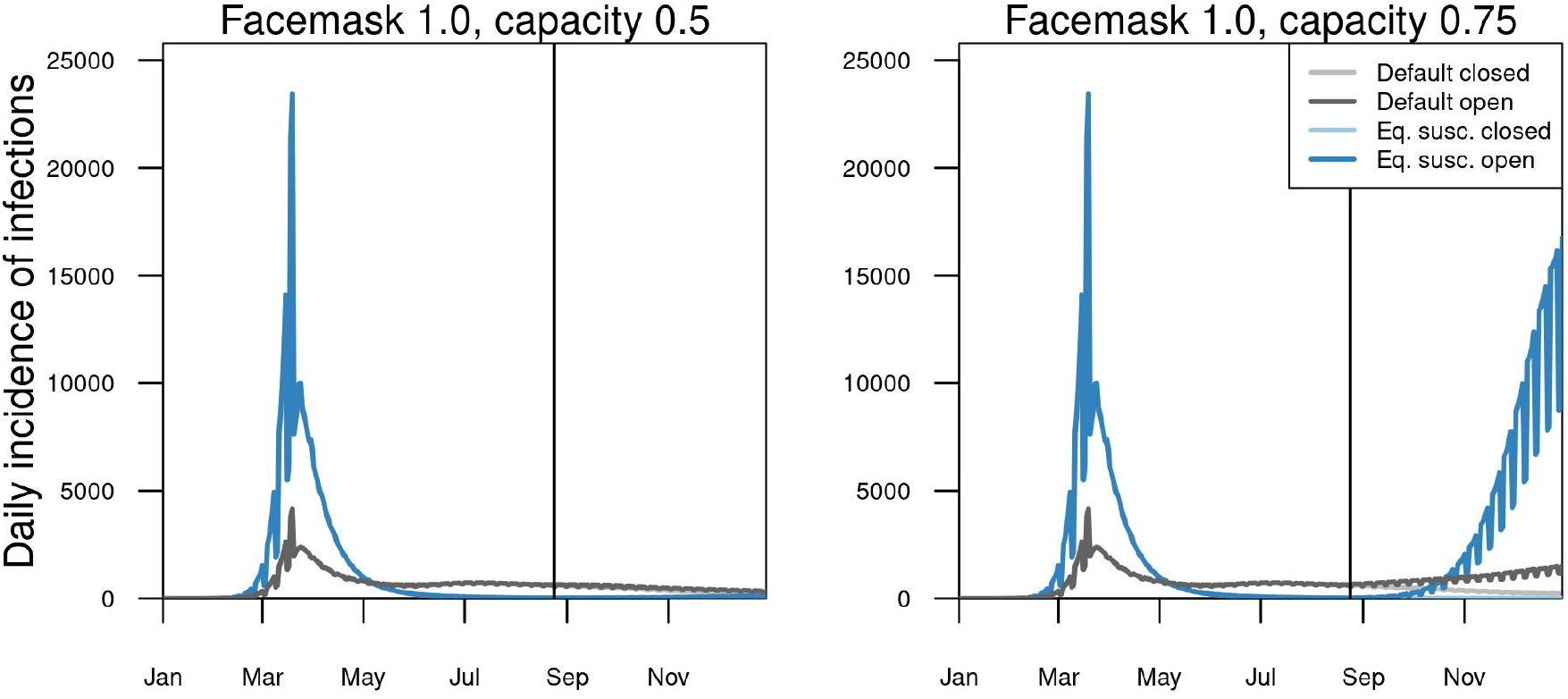
Comparison of median daily incidence of infection statewide in Indiana under alternative model assumptions and conditions in schools. Default model assumptions (gray) are contrasted with an alternative assumption of equal susceptibility for all ages (blue). A baseline scenario of schools operating remotely (light) is contrasted with two scenarios with schools reopened (dark): school operating capacity at 50% (left) or 75% (right). Face-mask adherence in schools was assumed to be 100% in both scenarios. The date on which schools reopened in the fall (August 24) is indicated by the vertical line.

**Figure S10.**
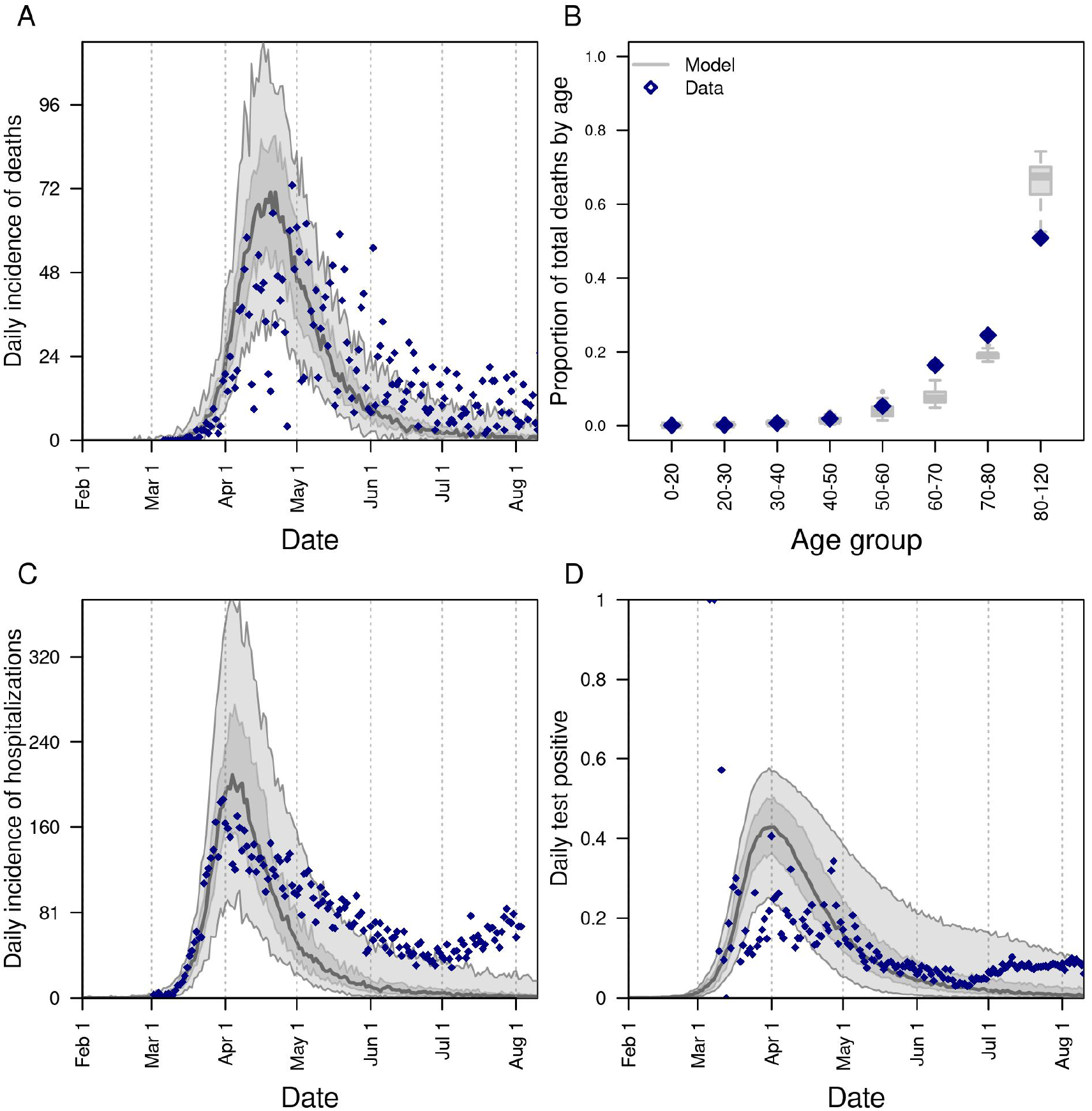
Model calibration to statewide data under an alternative assumption of equal susceptibility for all ages: A) daily incidence of death; B) proportion of deaths through July 13 in decadal age bins; C) daily incidence of hospitalization; and D) daily proportion of tests administered that are positive for SARS-CoV-2. In all panels, blue diamonds represent data. In A, C, and D, the gray line is the median, the dark shaded region the 50% posterior predictive interval, and the light shaded region the 95% posterior predictive interval.

**Table S1.** Proportional increase in risk of infection and death for the overall population of Indiana under alternative scenarios about school operating capacity and face-mask adherence in schools. We define the proportional increase in risk as the ratio of the cumulative number of events (infections or deaths) between August 24 and December 31 under the scenario indicated by the left two columns as compared to a scenario in which schools operate remotely.

| Capacity | Facemask | Cumulative infections | Cumulative deaths |
| --- | --- | --- | --- |
| 0.50 | 0.50 | 1.56 (1.48-1.64) | 1.19 (1.14-1.24) |
| 0.50 | 0.75 | 1.34 (1.28-1.41) | 1.13 (1.09-1.18) |
| 0.50 | 1.00 | 1.22 (1.16-1.28) | 1.09 (1.04-1.13) |
| 0.75 | 0.50 | 10.66 (10.06-11.28) | 2.64 (2.53-2.75) |
| 0.75 | 0.75 | 4.94 (4.63-5.28) | 1.81 (1.74-1.89) |
| 0.75 | 1.00 | 2.39 (2.26-2.52) | 1.39 (1.33-1.45) |
| 1.00 | 0.50 | 42.81 (41.44-44.44) | 9.22 (8.91-9.55) |
| 1.00 | 0.75 | 30.92 (29.82-32.10) | 6.18 (5.96-6.43) |
| 1.00 | 1.00 | 14.31 (13.58-15.10) | 3.30 (3.18-3.44) |

**Table S2.** Proportional increase in risk of infection and symptomatic infection for students under alternative scenarios about school operating capacity and face-mask adherence in schools. We define the proportional increase in risk as the ratio of the cumulative number of events (infections or symptomatic infections) between August 24 and December 31 under the scenario indicated by the left two columns as compared to a scenario in which schools operate remotely.

| Capacity | Facemask | Student infections | Student symptoms |
| --- | --- | --- | --- |
| 0.50 | 0.50 | 3.3 (3.1-3.5) | 3.1 (2.9-3.3) |
| 0.50 | 0.75 | 2.4 (2.3-2.5) | 2.3 (2.2-2.4) |
| 0.50 | 1.00 | 1.8 (1.7-1.9) | 1.8 (1.7-1.9) |
| 0.75 | 0.50 | 56.5 (53.1-60.1) | 49.0 (45.6-52.3) |
| 0.75 | 0.75 | 21.8 (20.3-23.6) | 18.9 (17.6-20.3) |
| 0.75 | 1.00 | 7.4 (6.9-7.9) | 6.6 (6.2-7.1) |
| 1.00 | 0.50 | 251.0 (241.2-260.9) | 236.4 (227.8-246.7) |
| 1.00 | 0.75 | 178.2 (170.1-186.1) | 162.8 (155.5-169.7) |
| 1.00 | 1.00 | 76.6 (72.1-81.3) | 67.7 (63.6-71.7) |

**Table S3.** Proportional increase in risk of infection, symptomatic infection, and death for teachers under alternative scenarios about school operating capacity and face-mask adherence in schools. We define the proportional increase in risk as the ratio of the cumulative number of events (infections, symptomatic infections, or deaths) between August 24 and December 31 under the scenario indicated by the left two columns as compared to a scenario in which schools operate remotely.

| Capacity | Facemask | Teacher infections | Teacher symptoms | Teacher deaths |
| --- | --- | --- | --- | --- |
| 0.50 | 0.50 | 2.1 (2.0-2.2) | 2.0 (1.9-2.2) | 2.0 (1.9-2.2) |
| 0.50 | 0.75 | 1.7 (1.6-1.8) | 1.7 (1.6-1.8) | 1.8 (1.7-2.0) |
| 0.50 | 1.00 | 1.5 (1.4-1.6) | 1.5 (1.4-1.6) | 1.6 (1.5-1.7) |
| 0.75 | 0.50 | 24.8 (23.2-26.5) | 23.6 (22.1-25.1) | 22.0 (20.2-23.7) |
| 0.75 | 0.75 | 10.7 (10.0-11.5) | 10.1 (9.4-10.9) | 9.8 (9.1-10.6) |
| 0.75 | 1.00 | 4.5 (4.2-4.8) | 4.4 (4.1-4.7) | 4.4 (4.1-4.7) |
| 1.00 | 0.50 | 119.5 (114.6-125.0) | 126.7 (120.8-132.9) | 166.1 (157.4-175.6) |
| 1.00 | 0.75 | 88.8 (84.9-93.1) | 90.2 (86.0-94.8) | 102.7 (96.6-109.6) |
| 1.00 | 1.00 | 41.8 (39.5-44.3) | 41.0 (38.5-43.7) | 41.1 (38.5-44.1) |

**Table S4.** Proportional increase in risk of infection, symptomatic infection, and death for family members of students and teachers under alternative scenarios about school operating capacity and face-mask adherence in schools. We define the proportional increase in risk as the ratio of the cumulative number of events (infections, symptomatic infections, or deaths) between August 24 and December 31 under the scenario indicated by the left two columns as compared to a scenario in which schools operate remotely.

| Capacity | Facemask | Family infections | Family symptoms | Family deaths |
| --- | --- | --- | --- | --- |
| 0.50 | 0.50 | 3.5 (3.3-3.7) | 3.3 (3.1-3.5) | 2.8 (2.6-3.0) |
| 0.50 | 0.75 | 2.5 (2.3-2.6) | 2.4 (2.2-2.5) | 2.3 (2.2-2.5) |
| 0.50 | 1.00 | 1.9 (1.8-2.0) | 1.9 (1.8-2.0) | 1.9 (1.7-2.0) |
| 0.75 | 0.50 | 55.8 (52.2-59.5) | 49.3 (46.0-52.5) | 32.5 (30.0-35.5) |
| 0.75 | 0.75 | 22.1 (20.5-23.6) | 19.4 (18.0-20.8) | 13.3 (12.3-14.4) |
| 0.75 | 1.00 | 7.7 (7.2-8.3) | 7.0 (6.6-7.6) | 5.7 (5.3-6.1) |
| 1.00 | 0.50 | 253.1 (242.4-263.4) | 249.1 (239.9-259.3) | 222.7 (211.2-236.5) |
| 1.00 | 0.75 | 177.7 (169.9-186.8) | 168.6 (161.4-176.2) | 132.0 (124.4-140.0) |
| 1.00 | 1.00 | 77.0 (72.6-81.7) | 69.5 (65.4-73.8) | 48.4 (44.8-52.2) |

**Table S5.** Risk ratio for the overall population of Indiana under alternative scenarios about school operating capacity and face-mask adherence in schools. We define the risk ratio as the ratio of the cumulative number of events (infections or deaths) between August 24 and December 31 under the scenario indicated by the left two columns as compared to a scenario in which schools operate at full capacity and without face masks.

| Capacity | Facemask | Cumulative infections | Cumulative deaths |
| --- | --- | --- | --- |
| 0.50 | 0.50 | 0.03 (0.03-0.03) | 0.09 (0.09-0.10) |
| 0.50 | 0.75 | 0.03 (0.03-0.03) | 0.09 (0.09-0.09) |
| 0.50 | 1.00 | 0.02 (0.02-0.02) | 0.08 (0.08-0.09) |
| 0.75 | 0.50 | 0.21 (0.20-0.22) | 0.21 (0.20-0.21) |
| 0.75 | 0.75 | 0.10 (0.09-0.10) | 0.14 (0.14-0.15) |
| 0.75 | 1.00 | 0.05 (0.04-0.05) | 0.11 (0.11-0.11) |
| 1.00 | 0.50 | 0.84 (0.83-0.85) | 0.72 (0.71-0.73) |
| 1.00 | 0.75 | 0.61 (0.60-0.62) | 0.48 (0.47-0.49) |
| 1.00 | 1.00 | 0.28 (0.27-0.29) | 0.26 (0.25-0.26) |

**Table S6.**
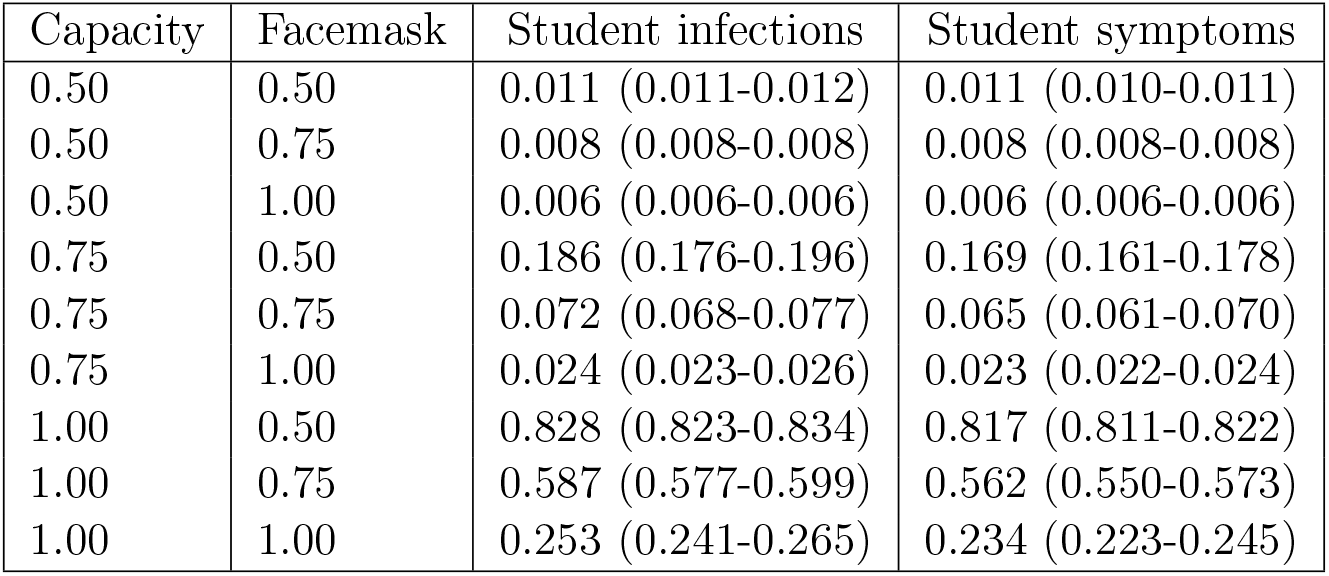
Risk ratio for students under alternative scenarios about school operating capacity and face-mask adherence in schools. We define the risk ratio as the ratio of the cumulative number of events (infections or symptomatic infections) between August 24 and December 31 under the scenario indicated by the left two columns as compared to a scenario in which schools operate at full capacity and without face masks.

**Table S7.** Risk ratio for teachers under alternative scenarios about school operating capacity and face-mask adherence in schools. We define the risk ratio as the ratio of the cumulative number of events (infections, symptomatic infections, or deaths) between August 24 and December 31 under the scenario indicated by the left two columns as compared to a scenario in which schools operate at full capacity and without face masks.

| Capacity | Facemask | Teacher infections | Teacher symptoms | Teacher deaths |
| --- | --- | --- | --- | --- |
| 0.50 | 0.50 | 0.015 (0.014-0.016) | 0.014 (0.013-0.014) | 0.009 (0.008-0.009) |
| 0.50 | 0.75 | 0.012 (0.012-0.013) | 0.011 (0.011-0.012) | 0.008 (0.008-0.008) |
| 0.50 | 1.00 | 0.011 (0.010-0.011) | 0.010 (0.010-0.010) | 0.007 (0.007-0.007) |
| 0.75 | 0.50 | 0.178 (0.170-0.187) | 0.157 (0.149-0.164) | 0.095 (0.090-0.100) |
| 0.75 | 0.75 | 0.077 (0.072-0.082) | 0.067 (0.063-0.071) | 0.043 (0.041-0.045) |
| 0.75 | 1.00 | 0.032 (0.031-0.034) | 0.029 (0.027-0.031) | 0.019 (0.018-0.020) |
| 1.00 | 0.50 | 0.858 (0.852-0.863) | 0.839 (0.832-0.844) | 0.723 (0.711-0.733) |
| 1.00 | 0.75 | 0.637 (0.626-0.648) | 0.598 (0.585-0.610) | 0.447 (0.433-0.461) |
| 1.00 | 1.00 | 0.300 (0.288-0.313) | 0.271 (0.259-0.283) | 0.179 (0.170-0.188) |

**Table S8.** Risk ratio for family members of students and teachers under alternative scenarios about school operating capacity and face-mask adherence in schools. We define the risk ratio as the ratio of the cumulative number of events (infections or deaths) between August 24 and December 31 under the scenario indicated by the left two columns as compared to a scenario in which schools operate at full capacity and without face masks.

| Capacity | Facemask | Family infections | Family symptoms | Family deaths |
| --- | --- | --- | --- | --- |
| 0.50 | 0.50 | 0.005 (0.005-0.005) | 0.005 (0.004-0.005) | 0.004 (0.004-0.005) |
| 0.50 | 0.75 | 0.003 (0.003-0.003) | 0.003 (0.003-0.003) | 0.003 (0.003-0.003) |
| 0.50 | 1.00 | 0.002 (0.002-0.002) | 0.002 (0.002-0.002) | 0.003 (0.003-0.003) |
| 0.75 | 0.50 | 0.081 (0.077-0.086) | 0.069 (0.066-0.072) | 0.042 (0.039-0.044) |
| 0.75 | 0.75 | 0.015 (0.014-0.015) | 0.013 (0.013-0.014) | 0.011 (0.011-0.012) |
| 0.75 | 1.00 | 0.004 (0.004-0.004) | 0.004 (0.004-0.004) | 0.004 (0.004-0.005) |
| 1.00 | 0.50 | 0.705 (0.694-0.716) | 0.673 (0.661-0.685) | 0.533 (0.519-0.546) |
| 1.00 | 0.75 | 0.241 (0.232-0.251) | 0.210 (0.201-0.218) | 0.128 (0.122-0.133) |
| 1.00 | 1.00 | 0.015 (0.015-0.016) | 0.014 (0.013-0.015) | 0.012 (0.012-0.013) |

**Table S9.** List of model parameters. The first nine were estimated through the model calibration process, and the subsequent fourteen were assumed based on the literature, as described in the Methods. Values of the first nine parameters reflect marginal posterior estimates.

| Parameter | Value |
| --- | --- |
| Importations scaling factor | 1.299 (95% CrI: 0.502-1.461) |
| Susceptibility slope | 0.757 (95% CrI: 0.578-3.344) |
| Susceptibility minimum | 0.346 (95% CrI: 0.311-0.506) |
| Susceptibility inflection point | 18.640 (95% CrI: 18.521-19.804) |
| Susceptibility maximum | 0.834 (95% CrI: 0.652-0.946) |
| Shelter in place adherence | 0.321 (95% CrI: 0.288-0.669) |
| Face-mask adherence | 0.534 (95% CrI: 0.461-0.540) |
| LTC importations scaling factor | 0.037 (95% CrI: 0.022-0.092) |
| Transmissibility | 0.593 (95% CrI: 0.501-0.788) |
| Probability of symptoms (by age) | Fig. S4A |
| Incubation period | Fig. S4B |
| Symptomatic period | Fig. S4C |
| Probability of death (by age) | Fig. S4D |
| Probability of death after symptom onset | Fig. S4E |
| Pre-infectious period | meanlog = 23.7, sdlog = 50 (Fig. S4F) |
| Infectious period | meanlog = 33.8, sdlog = 43.1 (Fig. S4F) |
| Adjusted odds ratio for face-mask efficacy | 0.30 [34] |
| School contacts | 0.6 [30, 36] |
| Classroom contacts | 1.2 [30, 36] |
| Workplace contacts | 0.06 [30, 36] |
| Office contacts | 0.13 [30, 36] |
| Household contacts | 0.14 [30, 36] |
| Neighborhood contacts | 0.78 [30, 36] |

**Table S10.**
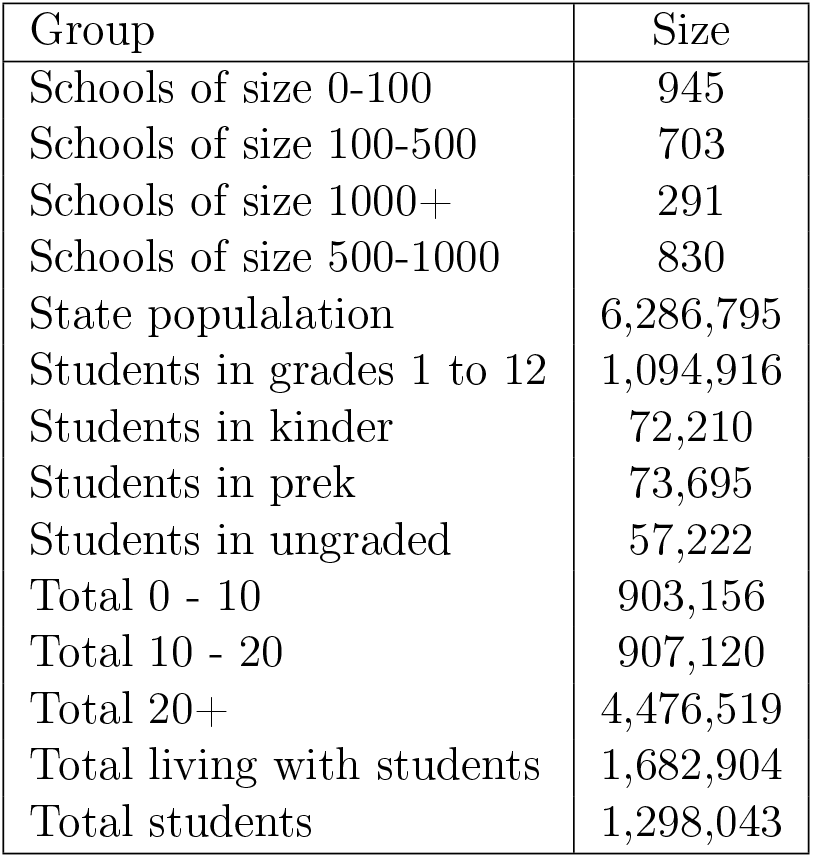
Summary of key features of the synthetic population of Indiana.

| Group | Size |
| --- | --- |
| Schools of size 0-100 | 945 |
| Schools of size 100-500 | 703 |
| Schools of size 1000+ | 291 |
| Schools of size 500-1000 | 830 |
| State populalation | 6,286,795 |
| Students in grades 1 to 12 | 1,094,916 |
| Students in kinder | 72,210 |
| Students in prek | 73,695 |
| Students in ungraded | 57,222 |
| Total 0 - 10 | 903,156 |
| Total 10 - 20 | 907,120 |
| Total 20+ | 4,476,519 |
| Total living with students | 1,682,904 |
| Total students | 1,298,043 |

**Table S11.**
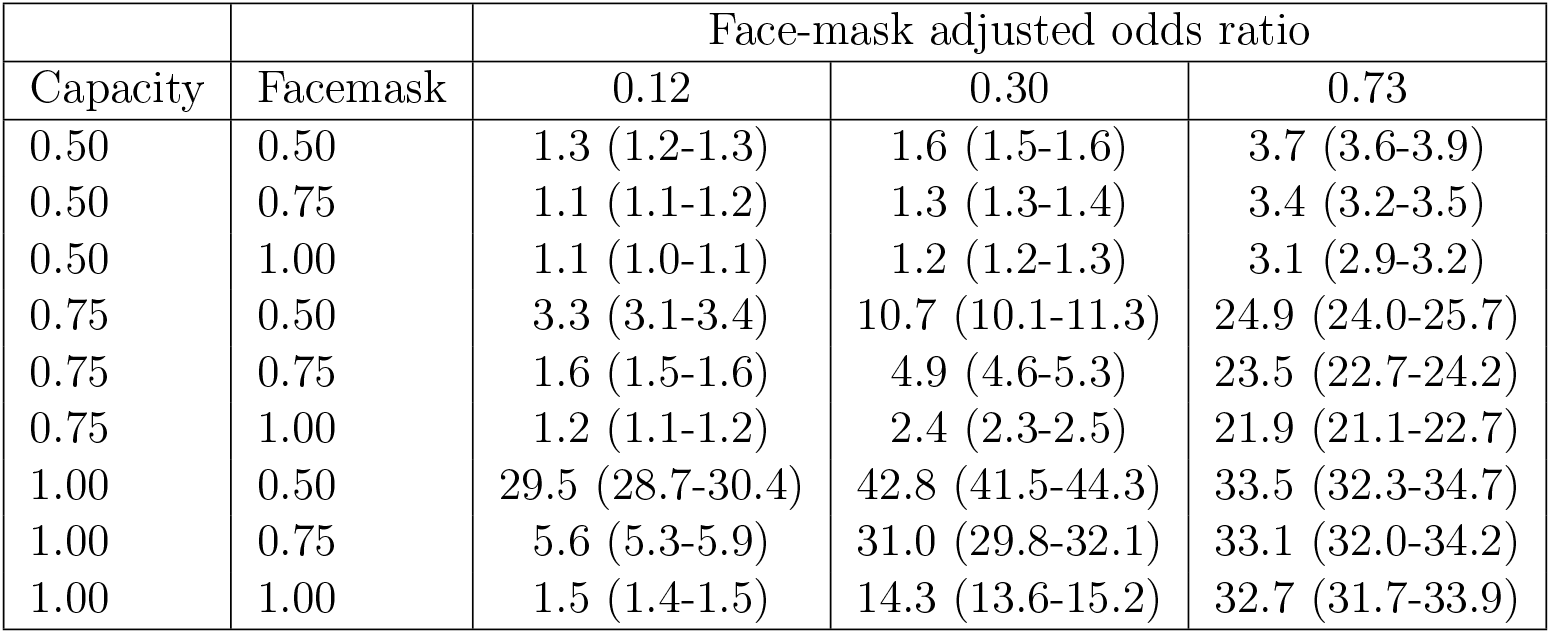
Proportional increases in cumulative infections statewide during fall 2020 under alternative values of face-mask protection, defined as the odds ratio (columns) and alternative scenarios about school operating capacity and face-mask adherence in schools (rows). These increases are relative to a scenario in which schools operated remotely in fall 2020.

|  |  | Face-mask adjusted odds ratio |  |  |
| --- | --- | --- | --- | --- |
| Capacity | Facemask | 0.12 | 0.30 | 0.73 |
| 0.50 | 0.50 | 1.3 (1.2-1.3) | 1.6 (1.5-1.6) | 3.7 (3.6-3.9) |
| 0.50 | 0.75 | 1.1 (1.1-1.2) | 1.3 (1.3-1.4) | 3.4 (3.2-3.5) |
| 0.50 | 1.00 | 1.1 (1.0-1.1) | 1.2 (1.2-1.3) | 3.1 (2.9-3.2) |
| 0.75 | 0.50 | 3.3 (3.1-3.4) | 10.7 (10.1-11.3) | 24.9 (24.0-25.7) |
| 0.75 | 0.75 | 1.6 (1.5-1.6) | 4.9 (4.6-5.3) | 23.5 (22.7-24.2) |
| 0.75 | 1.00 | 1.2 (1.1-1.2) | 2.4 (2.3-2.5) | 21.9 (21.1-22.7) |
| 1.00 | 0.50 | 29.5 (28.7-30.4) | 42.8 (41.5-44.3) | 33.5 (32.3-34.7) |
| 1.00 | 0.75 | 5.6 (5.3-5.9) | 31.0 (29.8-32.1) | 33.1 (32.0-34.2) |
| 1.00 | 1.00 | 1.5 (1.4-1.5) | 14.3 (13.6-15.2) | 32.7 (31.7-33.9) |

**Table S12.**
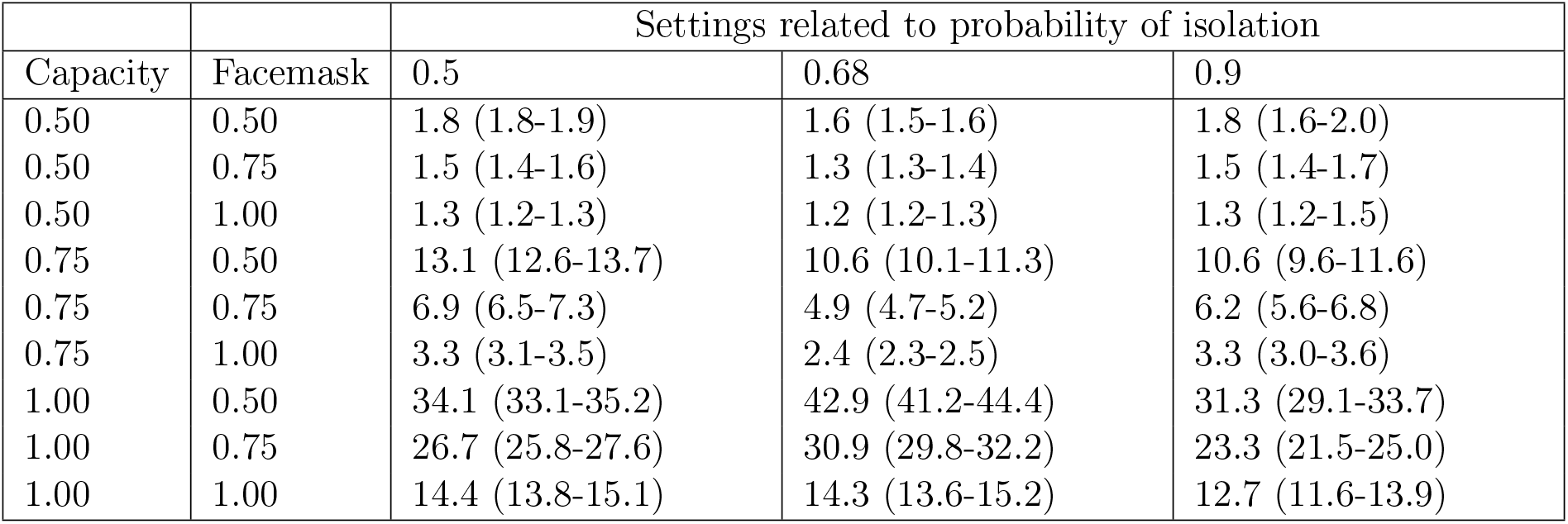
Proportional increases in cumulative infections statewide during fall 2020 under alternative values of the probability of isolation given symptoms (columns) and alternative scenarios about school operating capacity and face-mask adherence in schools (rows). These increases are relative to a scenario in which schools operated remotely in fall 2020.

|  |  | Settings related to probability of isolation |  |  |
| --- | --- | --- | --- | --- |
| Capacity | Facemask | 0.5 | 0.68 | 0.9 |
| 0.50 | 0.50 | 1.8 (1.8-1.9) | 1.6 (1.5-1.6) | 1.8 (1.6-2.0) |
| 0.50 | 0.75 | 1.5 (1.4-1.6) | 1.3 (1.3-1.4) | 1.5 (1.4-1.7) |
| 0.50 | 1.00 | 1.3 (1.2-1.3) | 1.2 (1.2-1.3) | 1.3 (1.2-1.5) |
| 0.75 | 0.50 | 13.1 (12.6-13.7) | 10.6 (10.1-11.3) | 10.6 (9.6-11.6) |
| 0.75 | 0.75 | 6.9 (6.5-7.3) | 4.9 (4.7-5.2) | 6.2 (5.6-6.8) |
| 0.75 | 1.00 | 3.3 (3.1-3.5) | 2.4 (2.3-2.5) | 3.3 (3.0-3.6) |
| 1.00 | 0.50 | 34.1 (33.1-35.2) | 42.9 (41.2-44.4) | 31.3 (29.1-33.7) |
| 1.00 | 0.75 | 26.7 (25.8-27.6) | 30.9 (29.8-32.2) | 23.3 (21.5-25.0) |
| 1.00 | 1.00 | 14.4 (13.8-15.1) | 14.3 (13.6-15.2) | 12.7 (11.6-13.9) |

**Table S13.** Proportional increases in cumulative infections statewide during fall 2020 under alternative assumptions about juvenile transmission (columns) and alternative scenarios about school operating capacity and face-mask adherence in schools (rows). These increases are relative to a scenario in which schools operated remotely in fall 2020.

| Capacity | Facemask | Settings related to juvenile transmission |  |  |  |
| --- | --- | --- | --- | --- | --- |
|  |  | Default | Equal Susc. | Low 0-10 Susc. | Low Asymp. Inf. |
| 0.50 | Low | 1.6 (1.5-1.6) | 6.1 (5.8-6.4) | 1.5 (1.4-1.6) | 1.7 (1.7-1.8) |
| 0.50 | Med | 1.3 (1.3-1.4) | 4.2 (4.0-4.4) | 1.3 (1.3-1.4) | 1.5 (1.4-1.5) |
| 0.50 | High | 1.2 (1.2-1.3) | 3.0 (2.9-3.1) | 1.2 (1.1-1.3) | 1.3 (1.3-1.4) |
| 0.75 | Low | 10.7 (10.0-11.3) | 348.0 (335.2-361.7) | 9.3 (8.8-9.8) | 10.0 (9.5-10.4) |
| 0.75 | Med | 4.9 (4.6-5.2) | 265.3 (254.7-276.8) | 4.6 (4.4-4.9) | 5.9 (5.7-6.2) |
| 0.75 | High | 2.4 (2.3-2.5) | 181.6 (174.0-189.9) | 2.3 (2.2-2.4) | 3.2 (3.0-3.3) |
| 1.00 | Low | 42.9 (41.4-44.5) | 687.5 (670.0-706.1) | 26.5 (25.4-27.7) | 26.2 (25.5-26.9) |
| 1.00 | Med | 31.0 (29.8-32.2) | 669.3 (652.3-686.8) | 20.8 (19.9-21.7) | 21.0 (20.3-21.6) |
| 1.00 | High | 14.3 (13.6-15.1) | 641.9 (624.5-659.7) | 11.3 (10.8-11.9) | 13.2 (12.7-13.8) |

## Supplementary methods

### Model description

#### People and their contacts

FRED simulates pathogen spread in a population by recreating interactions among people on a daily basis. To realistically represent the population of Indiana, we drew on a synthetic population of the US that represents demographic and geographic characteristics from 2010 [35]. Each human is modeled as an agent that visits a set of places defined by their activity space. This activity space contains places such as houses, schools, workplaces, and neighborhood locations. Transmission can occur when an infected person visits the same location as a susceptible person on the same day, with numbers of contacts per person specific to each location type. For instance, school contacts depend not on the size of the school but on the age of the student. We adopted contact rates specific to each location type that were previously calibrated to attack rates for influenza in each location type [30, 36].

#### Importation to seed local transmission

To initialize the model, we simulated international and domestic importations similar to Perkins et al. [57]. First, we obtained data on internationally and domestically imported deaths in Indiana up to March 18 [58], which we used to extrapolate total international and domestic importations based on the case fatality risk [38], the proportion of infections that are asymptomatic [37], and the probability of detecting local and international symptomatic infections [57]. Second, we assigned times to internationally imported infections proportional to international incidence patterns, adjusted to account for the timing of a ban on travel from China. We assigned times to domestically imported infections proportional to total US incidence. Drawing from uncertainty distributions for each of the three aforementioned parameters, we repeated this process 1,000 times and averaged across replicates. We used that average curve to seed our model, scaling its magnitude with a parameter that we calibrated. Although importations from outside Indiana likely continued beyond those that we were able to account for explicitly, we assumed that transmission within Indiana was sufficient at that point to be the primary driver of incidence. In addition to importations in the overall population, we simulated importation into long-term care facilities, given the large number of deaths that took place there and the limited realism of our model in simulating visitors to those facilities. We introduced infections into these facilities at a constant rate that we calibrated.

#### Transmission and disease progression

Once infected, each individual had latent and infectious periods drawn from distributions calibrated so that the average generation interval distribution matched estimates from Singapore (*µ* = 5.20, *σ* = 1.72) [33]. The absolute risk of transmission depended on the number and location of an infected individual’s contacts and a parameter that controls SARS-CoV-2 transmissibility upon contact, which we calibrated. We assumed asymptomatic infections were as infectious as symptomatic infections and had identical timing of infectiousness [39, 40, 26, 41]. Following exposure, we assumed that children were less susceptible to infection than adults, which we modeled with a modified logistic function calibrated to results of Davies et al. [19]. We defined four parameters of this function as the *minimum* susceptibility, the *maximum* susceptibility, the *inflection point* of susceptibility with respect to age, and the *slope* of the age-susceptibility relationship around the inflection point. For agents that developed symptoms, we took random draws from lognormal distributions for the incubation period [32] and duration of symptoms [31]. Both the probabilities of developing symptoms and dying [21] were assumed to increase with age. The probability of developing symptoms by age was estimated using the age distribution of cases in the China CDC report from early in the pandemic[38] and the age distribution of the population in China[79]. By assuming equal exposure by age and assuming that 50% of infections proceed asymptomatically (an intermediate value of two estimates from the Diamond Princess [37, 80]), we can calculate the proportion of symptomatic infections we would expect in each age group as under our assumptions, the total number of infections in an age-group will be proportional to the population size in that age-group. For infections that resulted in death, we modeled the time to death with a gamma distribution [22] truncated at the 99th percentile. These and other parameters are summarized in Table S9 and Fig. S4.

#### Changes in agent behavior during the epidemic

Agent behavior in FRED has the potential to change over the course of an epidemic. Following the onset of symptoms, infected agents self-isolate at home according to a fixed daily rate, whereas others continue their daily activities [42]. This rate is chosen so that on average 68% of agents will self-isolate at some point during their symptoms, assuming that all individuals who develop a fever will isolate at some point during their symptoms, based on published studies in the U.S. [43]. Agents can also respond to public health interventions, including school closure, shelter in place, and a combination of mask-wearing and social distancing. School closures occur on specific dates [81], resulting in students limiting their activity space to household and neighborhood locations. Shelter-in-place interventions reduce some agents’ activity spaces to their households only, whereas others continue with their daily routines. We used mobility reports from Google [46] to drive daily compliance with shelter-in-place, such that shelter-in-place compliance in our model accounts for both the effects of shelter-in-place orders and some people deciding to continue staying at home after those orders are lifted [47]. To account for voluntary mask-wearing and social distancing, we used Google Trends data for Indiana using the terms “face mask” and “social distancing” [48] and used estimates on face-mask adherence from a New York Times analysis of a survey from Dynata [49].

### Model calibration

We selected nine parameters to estimate based on calibration of the model to four data types on COVID-19 in Indiana: daily incidence of death, age distribution of deaths, daily incidence of hospitalization, and daily test positivity. The initial ranges for the statewide and long-term care facility importations were adjusted to cover a wide range of values. Compliance with shelter-in-place was informed with changes in mobility patterns in the Google community reports [46]. We fitted a GAM to the trends from the percentage change on mobility trends for places of residence, and projected the compliance of shelter-in-place orders after the period for which we had data by assuming a linear trend thereafter. We normalized these mobility trends from 0% (baseline) to 100% (everyone at home) and adjusted its magnitude with a parameter representing the maximum compliance in the historical trends. The minimum, maximum, inflection point, and slope of the logistic function with which we model the age-susceptibility relationship were calibrated to estimates by Davies et al. [19].

We simulated 6,000 combinations of these nine parameters, 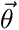, using a sobol design sampling algorithm with the sobolDesign function in R [51, 52]. For each parameter set, we calculated the likelihood of the model given the observed data on daily incidence of death, cumulative deaths in long-term care facilities through July 13, the decadal age distribution of cumulative deaths through July 13, daily incidence of hospitalization, and test positivity.

We calculated the contribution to the likelihood for daily incidence of death and cumulative deaths in long-term care facilities using a negative binomial distribution as

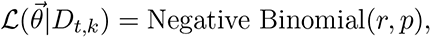

where *D_t,k_* is the daily incidence of death on day *t* and location *k* (long-term care facilities or all other locations), and *r* and *p* are size and probability parameters, respectively. We informed *r* and *p* using the conjugate prior relationship between a beta prior and negative binomial likelihood, such that *r* = *r_prior_* + *d_t,m_* and 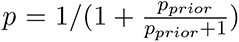, where *d_t,m_* is the daily incidence of death predicted by the model on day *t*. For the decadal age distribution of cumulative deaths through July 13, we used a multinomial distribution, such that

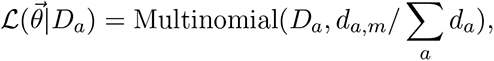

where *D_a_* is the observed number of deaths in the age group *a*, and *d_a,m_* are the deaths by age group obtained by the model. To fit to data on testing, we first observe, using Bayes’ rule, that

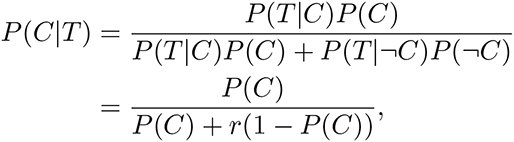

where *C* refers to a symptomatic case, *T* refers to an administered PCR test for current infection, and 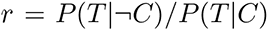. Next, we assume that non-symptomatic infections (either presymptomatic or asymptomatic) exhibit treatment-seeking behavior similar to uninfected individuals, or 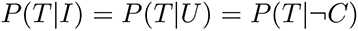, where *I* refers to a non-symptomatic infection and *U* to uninfected. We then observe, again using Bayes’ rule, that

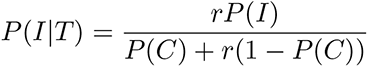

and

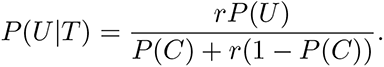

Next, we incorporate PCR sensitivity and specificity by assuming that sensitivity = (*P|C*) = *P* (*P|I*), where *P* refers to a positive test (i.e., we assume that PCR sensitivity is similar for non-symptomatic and symptomatic infections). This allows us to write

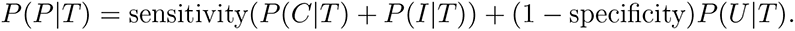

Then, we are in a position to write the contribution to the likelihood from the testing data, assuming that the number of positive tests in the data, *T*_+_, follows a binomial distribution

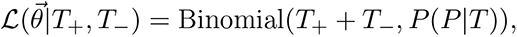

where *T_−_* represents the number of negative tests in the data.

Finally, the combined log-likelihood was obtained as

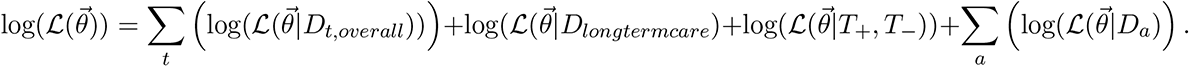

We sampled the parameters proportional to their likelihood to obtain a set of parameter combinations that constitute our approximation of the posterior distribution of parameter values.

## Notes

### Competing Interest Statement

The authors have declared no competing interest.

### Author Declarations

This is a simulation model and does not require IRB approval

### Summary of Updates

We have significantly revised the methods section to make the organization clearer. We have changed the simulation experiments to analyze different scenarios in the past with different conditions in schools. We also extended the model calibration to include fall 2020.

